# Variant-Level Functional Classification of Monoallelic TP53 Mutations Refines Prognostic Stratification in Myelodysplastic Neoplasms Beyond Allelic Status

**DOI:** 10.64898/2026.03.18.26348425

**Authors:** Alexander Streuer, Yotaro Ochi, Vladimir Riabov, Yasuhito Nannya, Laurenz Steiner, Mohammed Abba, Georgia Metzgeroth, Eva Altrock, Felicitas Rapp, Verena Nowak, Edin Hepgülüm, Daniel Nowak, Wolf-Karsten Hofmann, Seishi Ogawa, Nanni Schmitt

**Affiliations:** Department of Hematology and Oncology, Medical Faculty Mannheim, Heidelberg University, Mannheim, Germany; Department of Pathology and Tumor Biology, Graduate School of Medicine, Kyoto University, Kyoto, Japan; Division of Hematopoietic Disease Control, The Institute of Medical Science, The University of Tokyo, Tokyo, Japan; Faculty of Biosciences, Heidelberg University, Heidelberg, Germany; Institute for the Advanced Study of Human Biology (WPI-ASHBi), Kyoto University, Kyoto, Japan; Department of Innovative Medicine, Faculty of Medicine, Kindai University, Osaka, Japan

## Abstract

*TP53* mutations represent one of the strongest adverse prognostic factors in myelodysplastic neoplasms (MDS). While multi-hit *TP53* (*TP53^multiHit^*) alterations uniformly lead to very poor outcomes, the prognostic relevance of monoallelic *TP53* (*TP53^mono^*) mutations remains controversial. *TP53* variants can cause loss-of-function, dominant-negative, or gain-of-function effects. We hypothesized that functional heterogeneity among *TP53* variants contributes to the variable clinical behavior observed in monoallelic *TP53*-mutated MDS. Therefore, we analyzed pretreatment samples from 4,505 patients with MDS from two independent cohorts (IWG, n=3,173; J-MDS, n=1,332), including 271 patients with *TP53^mono^* and 499 with *TP53^multiHit^*. Functional annotation of *TP53* variants was performed using a previously published phenotype score (PS) derived from saturation mutagenesis screens, capturing dominant-negative and loss-of-function effects.

Median overall survival (OS) differed significantly by *TP53* allelic state (*TP53* wild-type (*TP53^wt^*) 42.4 months; *TP53^mono^* 22.9 months; *TP53^multiHit^* 9.2 months; p < 0.001). Within the *TP53^mono^* subgroup, functional annotation identified marked heterogeneity. Patients with high PS (≥7) showed significantly inferior OS compared with those with low PS (median OS: 13.8 vs. 39.2 months; HR 1.68, 95% CI 1.16-2.42; p = 0.006), particularly for IPSS-R and IPSS-M low-risk cases. Combining PS and variant allele frequency (VAF) further improved risk stratification. *TP53^mono^* patients with PS ≥7 and VAF ≥22% had outcomes comparable to *TP53^multiHit^* (median OS: 8.8, p = 0.2), whereas those with PS <7 and VAF <22% exhibited survival similar to *TP53^wt^*(median OS: 49.7, p = 0.9).

Overall, functional annotation of *TP53* variants refines prognostication in *TP53^mono^*-mutated MDS and may enhance individualized risk assessment.

## Introduction

Myelodysplastic neoplasms (MDS) are a group of clinically and genetically heterogeneous hematological disorders [1]. Among the various genetic abnormalities in MDS, mutations in the *TP53* gene have emerged as among the most significant negative prognostic factors [2–7].

Several studies have suggested that higher *TP53* variant allele frequency (VAF) correlates with worse overall survival (OS) in *TP53*-mutated MDS, although the thresholds and the independent prognostic value of VAF vary between cohorts and analyses [6, 8, 9].

Notably, the prognostic impact of monoallelic *TP53* (*TP53^mono^*) mutations appears to vary considerably across studies. In a large cohort analysis of n=368 *TP53*-mutated MDS patients, *TP53^mono^* mutations were associated with outcomes largely comparable to *TP53* wild-type (*TP53^wt^*) MDS [5], whereas the adverse prognostic effect was primarily driven by multi-hit *TP53* (*TP53^multiHit^*) alterations. Consistently, *TP53^multiHit^* status is the strongest adverse molecular prognostic factor for OS in the IPSS-M, whereas monoallelic *TP53* mutations did not confer additional prognostic information and were therefore not incorporated into the score [2]. In contrast, in another cohort of n=155 *TP53*-mutated MDS patients, patients with *TP53^mono^* mutations exhibited survival outcomes much closer to those of *TP53^multiHit^* cases than to *TP53^wt^* patients [10]. These discrepancies indicate substantial biological heterogeneity within *TP53^mono^*-mutated MDS.

The *TP53* gene encodes the p53 tumor suppressor protein, a transcription factor essential for cellular stress responses, such as DNA damage and oncogenic activation [11]. Efficient sequence-specific DNA binding and transcriptional activation require the assembly of four functionally competent p53 subunits into a tetramer, whereas incorporation of dysfunctional subunits compromises transcriptional activity [12, 13]. Upon activation, p53 induces a broad array of anti-tumor processes, including cell cycle arrest, DNA repair, apoptosis and senescence modulation of stem cell fate decisions [14–16].

In MDS, *TP53* is predominantly affected by missense mutations within the DNA-binding domain, which impair sequence-specific transcriptional activation of canonical p53 target genes [17, 18]. Many *TP53* missense variants result in loss of transcriptional function, either by disrupting DNA-contact residues or by inducing conformational instability of the p53 protein, thereby preventing effective promoter binding [15, 19]. In addition, mutant p53 proteins frequently retain the tetramerization domain, enabling the formation of hetero-tetramers with wild-type p53 and exerting dominant-negative effects, whereby incorporation of mutant subunits renders the entire p53 tetramer dysfunctional, even when only one *TP53* allele is affected [3, 12].

Beyond functional inactivation, some *TP53* mutants acquire gain-of-function properties, actively promoting oncogenic phenotypes through altered transcriptional programs [20, 21].

To systematically capture the functional consequences of individual *TP53* variants, Giacomelli et al. performed comprehensive saturation mutagenesis screens across all possible *TP53* missense mutations, integrating loss-of-function and dominant-negative effects in cellular fitness assays [22]. This approach enabled the derivation of a quantitative *TP53* phenotype score (PS), reflecting the extent to which individual variants attenuate canonical p53 transcriptional activity.

Despite these insights, current clinical risk stratification largely treats *TP53* mutations as a binary variable or relies on allelic state, without systematically accounting for variant-specific functional effects. A quantitative framework integrating *TP53* mutation-specific functional impact with clinical outcome data is therefore needed to refine prognostication in *TP53*-mutated MDS.

In this study, we analyzed large, well-annotated international cohorts together with previously published high-throughput functional data to investigate whether a *TP53* PS capturing transcriptional loss-of-function and dominant-negative activity can explain the prognostic heterogeneity observed in *TP53^mono^*-mutated MDS and improve individualized risk assessment.

## Methods

### Study Cohorts and Patient Selection

A total of 4,505 patients with a diagnosis of MDS and available comprehensive molecular profiling and survival data were included in this study. The dataset consisted of 3,173 patients from the international IWG cohort and an additional 1,332 patients from the independent Japanese J-MDS cohort. The IWG cohort dataset was retrieved from cBioPortal for Cancer Genomics (https://www.cbioportal.org), including clinical annotations, sequencing results, and mutation calls for each patient. Only cases with survival data were included.

The J-MDS cohort was generated using targeted next-generation sequencing of pretreatment samples. Single-nucleotide variants and small insertions/deletions were called using a variant allele frequency threshold of 2.5%. *KMT2A* (*MLL*) partial tandem duplications were detected either through dedicated structural variant calling using the genomonSV [23] pipeline or by panel-specific detection algorithms. MDS diagnoses were established according to contemporaneous WHO criteria at the respective institutions.

OS was defined as the interval from the date of initial diagnosis to death or last follow-up. All analyses were conducted on anonymized datasets in accordance with institutional and national ethical regulations.

### Molecular Profiling and Definition of TP53 Allelic States

In the IWG dataset, putative oncogenic variants had been pre-classified as pathogenic as previously described using a multi-database rule-based annotation framework [5]. For the Japanese cohort, the same criteria were applied.

*TP53* allelic state was assigned as previously described [2, 5], with multi-hit status defined by the presence of multiple *TP53* point mutations, or a *TP53* mutation accompanied by a del(17p) detected by conventional cytogenetics, or by loss of heterozygosity (LOH), including copy-number neutral LOH, identified through CNACS algorithm [24].

### Functional Annotation of TP53 Variants

Functional annotation of *TP53* variants was performed using a PS as previously described by Giacomelli et al. [22]. This PS is derived from comprehensive saturation mutagenesis screens assessing the functional impact of all possible *TP53* missense and nonsense variants in an isogenic wild-type p53 and p53NULL A549 human lung carcinoma cell line model.

Briefly, pooled *TP53* variant libraries were lentivirally transduced into isogenic wild-type p53 and p53NULL cells and subjected to positive and negative selection assays under p53-activating conditions. Variant-specific enrichment or depletion was quantified using normalized Z-scores. For each transduced *TP53* variant, the PS was calculated as the mean of Z-scores obtained from three experimental conditions: (a) wild-type p53 cells treated with nutlin-3, a p53-MDM2 inhibitor, reflecting dominant-negative activity of the transduced variant through interference with endogenous wild-type p53; (b) p53NULL cells treated with nutlin-3, capturing loss-of-function effects of the transduced variant in the absence of wild-type p53; and (c) p53NULL cells treated with etoposide, a topoisomerase inhibitor, assessing wild-type-like p53 activity of the transduced variant under DNA damage-induced stress.

Higher PS values reflect stronger dominant-negative or loss-of-function effects on p53 activity, whereas lower PS values indicate preserved or wild-type-like function.

### Statistical Analysis

Survival curves were estimated using the Kaplan-Meier method and compared between groups using the log-rank test.

Univariable and multivariable Cox proportional hazards regression models were applied to assess the prognostic impact of analyzed factors.

Continuous variables were compared using the Wilcoxon rank-sum test and categorical variables were compared using Fisher’s exact test. Correlations between continuous variables were assessed using Spearman’s rank correlation coefficient.

All statistical tests were two-sided, and a p value < 0.05 was considered statistically significant. Statistical analyses were performed using R (version 4.3.1), with survival analyses conducted using the *survival* (v3.5.5) and *survminer* (v0.5.0) packages.

## Results

### Influence of allelic state of TP53 on survival of MDS patients

In this study, a total of 4,505 patients with complete molecular profiling covering all IPSS-M relevant genes were analyzed, including 271 *TP53^mono^* and 499 *TP53^multiHit^*patients.

*TP53* mutational status exerted a significant impact on OS, as previously described. Depending on the allelic status, median OS was 42.4 months for patients with *TP53^wt^*, 22.9 months for *TP53^mono^*, and 9.2 months for *TP53^multiHit^* (p < 0.001; **Figure 1A**). In the IWG cohort, the difference in OS between *TP53^wt^* and *TP53^mono^*was modest and did not reach statistical significance (median OS: 43.1 vs. 29.7 months; HR 1.22, 95% CI: 0.99-1.52; p = 0.07; **Supplementary Figure 1A**), whereas the J-MDS cohort demonstrated a substantially worse survival outcome for the *TP53^mono^* patients (median OS: 36.7 vs. 12.3 months; HR 2.65, 95% CI: 2.03-3.47; p < 0.001; **Supplementary Figure 1B**). To identify plausible causes for this cohort-specific divergence, additional comparative analyses were conducted.

**Figure 1.**
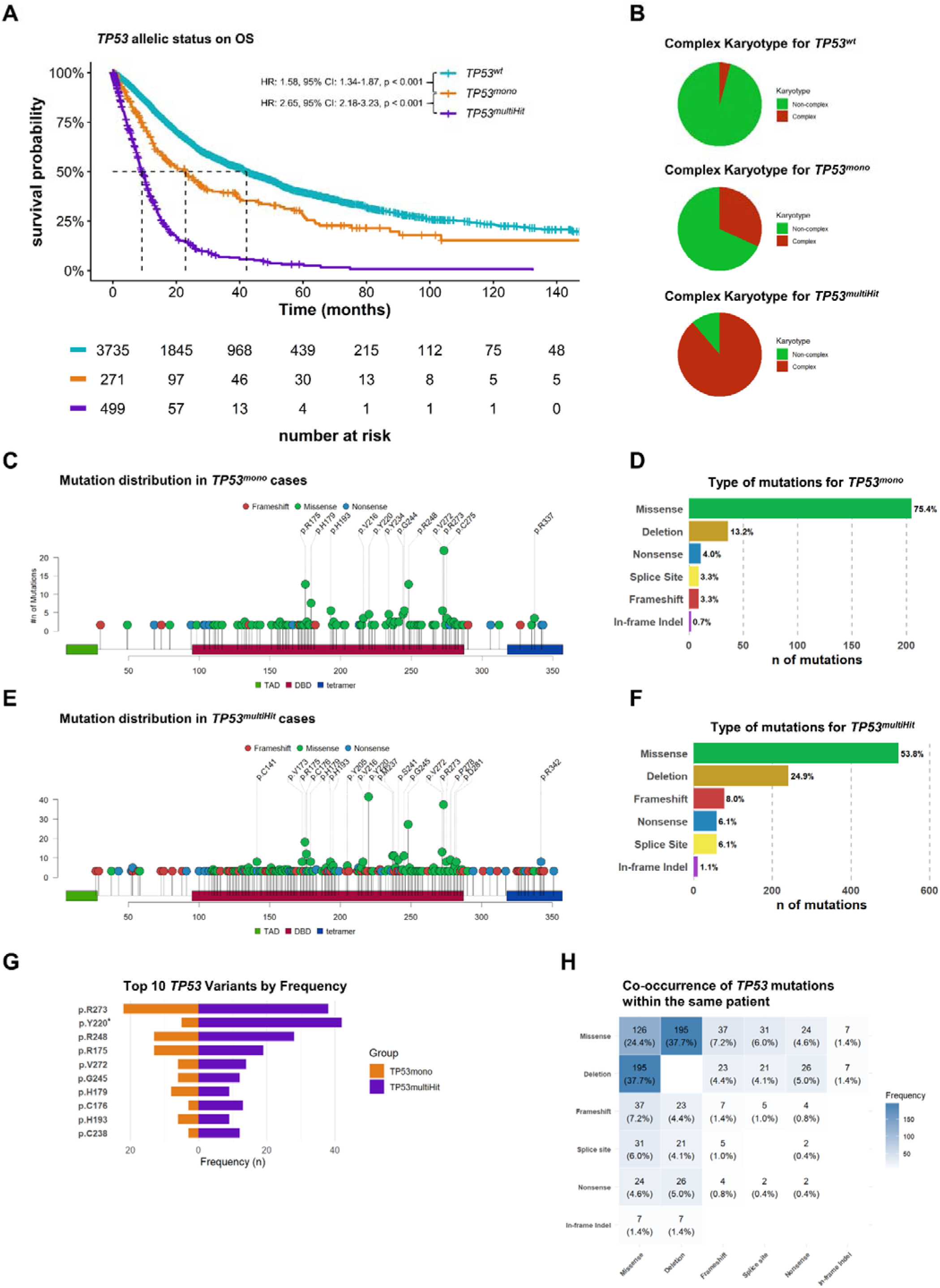
*TP53* allelic state defines survival, karyotype complexity, and mutational landscape in MDS. (A) Kaplan-Meier estimates of overall survival (OS) stratified by *TP53* allelic state (*TP53^wt^*, *TP53^mono^*, *TP53^multiHit^*). *TP53^mono^* cases show inferior survival compared with *TP53^wt^*, while *TP53^multiHit^*cases exhibit the poorest outcomes. (B) Proportion of complex versus non-complex karyotypes across *TP53* allelic states, demonstrating a progressive increase in karyotype complexity from *TP53^wt^* to *TP53^mono^*and *TP53^multiHit^*. (C) Positional distribution of *TP53* mutations across the protein in *TP53^mono^* cases, showing predominant clustering within the DNA-binding domain (DBD; 90.1% of variants). (D) Distribution of *TP53* mutation types in *TP53^mono^*cases, highlighting the predominance of missense variants (75.4%). The category “deletion” denotes cytogenetic *TP53* aberrations, including 17p deletion/copy-number loss and cnLOH. (E) Positional distribution of *TP53* mutations in *TP53^multiHit^* cases, demonstrating a broader distribution across the protein with a reduced proportion of variants located within the DBD (80.7% vs 91.4% in *TP53^mono^*; p < 0.001). (F) Distribution of *TP53* mutation types in *TP53^multiHit^*cases, showing increased proportions of truncating and structural alterations compared with *TP53^mono^* (deletions: 23.1% vs 13.2%; frameshift: 8.3% vs 3.3%; stop-gain: 5.7% vs 4.0%). Notably, 96.1% of missense variants in *TP53^multiHit^* cases remained located within the DBD. (G) Top ten most frequent *TP53* variants stratified by allelic state (*TP53^mono^* vs *TP53^multiHit^*). Asterisk (*) indicates statistically significant enrichment between allelic states. (H) Co-occurrence patterns of *TP53* mutation classes within individual patients, illustrating the frequency of different mutation-type combinations in *TP53^multiHit^* cases.

Furthermore, a complex karyotype strongly correlated with *TP53* allelic status. It was rare in *TP53^wt^* patients (4.3%, 160/3735), present in 32.8% of *TP53^mono^* cases (89/271), and observed in the vast majority of *TP53^multiHit^* patients (88.8%, 443/499; p < 0.001; **Figure 1B**), consistent with prior reports [6, 10, 25].

We next investigated the distribution of *TP53* mutations across the protein structure. In line with previous findings [17, 26, 27], mutations were predominantly located within the DNA-binding domain (DBD), suggesting a non-random distribution. In the monoallelic context, the majority of mutations (90.1%) were located within the DBD (**Figure 1C**), with missense mutations representing the predominant type (75.4%, **Figure 1D**).

However, the mutational landscape shifts notably in the multi-hit state, as a strong increase in truncating mutations was observed (**Figures 1E–F**).

The ten most frequent *TP53* mutations are listed in **Table 1** for *TP53^mono^* and in **Table 2** for *TP53^multiHit^*, while the complete lists of all detected variants are provided in **Supplementary Table 1** and **Supplementary Table 2**, respectively.

**Table 1.** Top 10 most frequent variants in *TP53^mono^*.

| **Protein Change** | **Protein Position** | **Variant Classification** | **No. of cases** |
| --- | --- | --- | --- |
| p.R273 | 273 | Missense | 22 (9.6%) |
| p.R175 | 175 | Missense | 13 (5.7%) |
| p.R248 | 248 | Missense | 13 (5.7%) |
| p.H179 | 179 | Missense | 8 (3.5%) |
| p.H193 | 193 | Missense | 6 (2.6%) |
| p.G245 | 245 | Missense | 6 (2.6%) |
| p.V272 | 272 | Missense | 6 (2.6%) |
| p.Y220 | 220 | Missense | 5 (2.2%) |
| p.Y234 | 234 | Missense | 5 (2.2%) |
| p.G244 | 244 | Missense | 5 (2.2%) |

**Table 2.** Top 10 most frequent *TP53* variants in *TP53^multiHit^*.

| **Protein Change** | **Protein Position** | **Variant Classification** | **No. of cases** |
| --- | --- | --- | --- |
| p.Y220 | 220 | Missense | 42 (6.0%) |
| p.R273 | 273 | Missense | 38 (5.4%) |
| p.R248 | 248 | Missense | 28 (4%) |
| p.R175 | 175 | Missense | 19 (2.7%) |
| p.V272 | 272 | Missense | 14 (2.0%) |
| p.C176 | 176 | Missense | 13 (1.9%) |
| p.M237 | 237 | Missense | 12 (1.7%) |
| p.C238 | 238 | Missense | 12 (1.7%) |
| p.G245 | 245 | Missense | 12 (1.7%) |
| p.S241 | 241 | Missense | 10 (1.4%) |
| p.P278 | 278 | Missense | 10 (1.4%) |

In the monoallelic state, the most frequent variants were *p.R273* (n=22, 9.6%), *p.R175* (n=13, 5.7%) and *p.R248* (n=13, 5.7%) whereas in the multi-hit state, *p.Y220* (n=42, 6.0%) and *p.R273* (n=38, 5.4%) were predominantly present (**Figure 1G**).

Among all variants, only one was significantly enriched in multi-hit cases compared to monoallelic ones*: p.Y220* (5 (2.2%) vs. 42 (6.0%), p = 0.007, **Figure 1G**), indicating a preferential association with biallelic *TP53* states.

Additionally, we analyzed co-mutation patterns among multi-hit cases. Combinations involving two missense mutations accounted for only 24.4% of all observed *TP53* co-mutations. The remaining consisted of combinations involving at least one truncating or structural variant (**Figure 1H**).

### The phenotype of TP53 mutations has a strong impact on overall survival in the mono-allelic state

To account for the functional diversity of *TP53* mutations, each variant was functionally characterized. Functional annotation of all possible *TP53* variants was based on published high-throughput saturation mutagenesis data obtained through a pooled screening approach in a human cell line. Variants were classified as loss-of-function or dominant-negative effect according to their enrichment under selective conditions. A PS was derived from individual enrichment values to standardize the interpretation of each allele’s functional impact [22]. The PS reflects the functional pathogenicity of individual *TP53* variants, with higher values indicating stronger dominant-negative or loss-of-function effects and lower values indicating near wild-type activity. This PS was subsequently applied to assess the prognostic impact of *TP53* variants in MDS patients.

For the *TP53^mono^* cohort, the PS ranged from 0.20 (*TP53 p.R342\**) to 21.30 (*TP53 p.H179Q*) with a median of 6.8 and an interquartile range of 4.7-10.6 (**Supplementary Figure 2A, B**). When plotting continuous PS cutoffs against OS, we observed that higher PS values were associated with inferior survival, whereas lower PS values indicated better outcomes, with an optimal cutoff around 7-10 (**Supplementary Figure 3**). Based on these findings, we selected a PS cutoff of 7, which was close to the cohort median. MDS patients harboring a *TP53^mono^* mutation with a high PS (≥7; n=110) had a significantly shorter OS compared to those with a low PS (<7; n=105; median OS: 13.8 vs 39.2; HR 1.68, 95% CI: 1.16-2.42; p = 0.006, **Figure 2A**). This difference was observed in both cohorts. As noted above, *TP53^mono^*-mutated patients in the J-MDS cohort had markedly shorter OS compared to those in the IWG cohort. Within each cohort, the PS separated monoallelic cases into subgroups with better and worse outcomes, although statistical significance was narrowly not reached because of the smaller subset (IWG: HR 1.30, 95% CI: 0.8-2.1; p = 0.3; J-MDS: HR 1.66, 95% CI: 0.9-2.9; p = 0.08; **Supplementary Figure 4A, B**). However, when excluding cases with a complex karyotype in the J-MDS cohort, the separation became very pronounced (median OS: 65.2 vs. 8.3 months; HR 3.1, 95% CI: 1.1-8.4; p = 0.03; **Supplementary Figure 4D**).

**Figure 2.**
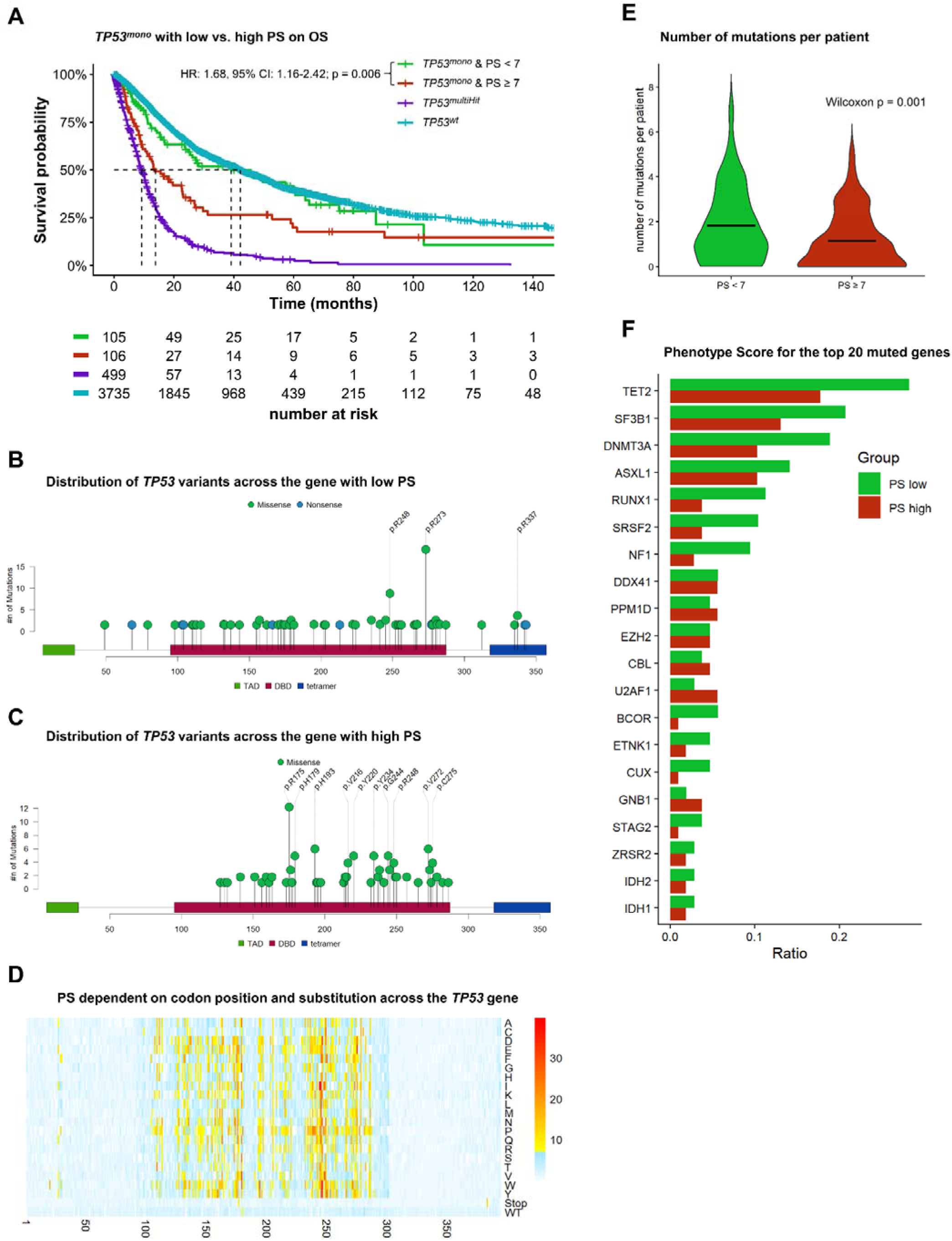
Phenotype Score based stratification of *TP53^mono^* mutations and their impact on survival and protein distribution. (A) Kaplan-Meier estimates of overall survival (OS) for *TP53^mono^* patients stratified by low (< 7) and high (≥ 7) phenotype score (PS), shown in the context of *TP53^wt^*and *TP53^multiHit^* reference groups. High PS *TP53^mono^* cases exhibit significantly inferior OS compared with low PS *TP53^mono^* cases. (B) Positional distribution of *TP53* variants with low PS (< 7) across the *TP53* protein (C) Positional distribution of *TP53* variants with high PS (≥ 7) across the *TP53* protein, demonstrating clustering within the DNA-binding domain (DBD). (D) Heatmap showing PS values across the *TP53* gene according to codon position and amino acid substitution, illustrating the relationship between mutation position, substitution type, and functional severity. (E) Patients with PS < 7 show a higher number of co-mutations compared with PS ≥ 7. (F) Frequency of recurrent co-mutated genes stratified by PS group.

To assess whether survival differences between the IWG and J-MDS cohorts were associated with differences in *TP53* PS distribution, we compared PS values across cohorts. PS values were significantly lower in the IWG cohort than in J-MDS (mean 6.55 vs. 8.32, p < 0.001; **Supplementary Figure 5A**).

Similarly, *TP53^mono^* cases in the IWG cohort also showed lower mean *TP53* VAF levels compared to the J-MDS cohort (mean VAF 21% vs. 27%, p = 0.002, **Supplementary Figure 5B**).

Analysis of the most frequent *TP53* variants revealed no significant differences in individual variant frequencies between IWG and J-MDS. Instead, cohort differences in PS and VAF reflected a global shift toward higher values in J-MDS rather than enrichment of specific variants (**Supplementary Figure 5C**).

Consistent with these differences, the J-MDS cohort also showed a higher proportion of *TP53*-mutated cases, affecting both *TP53^mono^* (8% vs. 5%, p < 0.001) and *TP53^multiHit^* disease (19% vs. 8%; p < 0.001; **Supplementary Figure 5D**).

Notably, PS provided additional prognostic resolution especially within lower-risk categories by identifying patients with adverse outcomes despite favorable baseline risk. Among IPSS-R low-risk patients, median OS was 63.9 months for PS <7 versus 23.1 months for PS ≥7 (HR 2.07, 95% CI 1.17–3.65; p = 0.01; **Supplementary Figure 6A**). Similarly, within IPSS-M low-risk disease, median OS was 63.9 versus 23.1 months for low versus high PS (HR 1.98, 95% CI 1.02–3.86; p = 0.04; **Supplementary Figure 6C**).

Beyond OS, higher *TP53* PS was also associated with increased leukemic transformation risk. *TP53^mono^* patients with PS ≥7 had a higher incidence of acute myeloid leukemia (AML) progression compared with lower PS (HR 1.93, 95% CI 1.0-3.7; p = 0.05; **Supplementary Figure 7A**), with similar trends in both cohorts. Although J-MDS showed a clear curve separation, statistical significance was not reached in cohort-specific analyses (**Supplementary Figure 7B, C**). A higher cutoff (PS ≥ 10) strongly predicted transformation in the IWG cohort (HR 3.29, 95% CI 1.1–9.7; p = 0.02) but was too stringent to effectively discriminate risk in J-MDS (**Supplementary Figure 7D-F**).

We next analyzed the distribution of low and high PS *TP53* variants across the gene (**Figure 2B-C**; **Tables 3-4**). Most recurrent hotspot variants clustered within the PS ≥7 group. Notable exceptions were *p.R273H* (PS 4.74) and *p.R248Q* (PS 6.24), which were associated with more favorable OS compared with hotspot high PS variants (*p.R175H*, *p.V272M*; **Supplementary Figure 8A**).

**Table 3.** Most frequent *TP53* variants in *TP53^mono^* with PS < 7.

| **Protein Change** | **Protein Position** | **Variant Classification** | **Phenotypic Score** | **No. of cases** |
| --- | --- | --- | --- | --- |
| p.R273H | 273 | Missense | 4.74 | 12 (11.1%) |
| p.R248Q | 248 | Missense | 6.24 | 9 (8.3%) |
| p.R273C | 273 | Missense | 6.43 | 7 (6.5%) |
| p.V157F | 157 | Missense | 3.59 | 3 (2.8%) |
| p.G245S | 245 | Missense | 5.65 | 3 (2.8%) |
| p.V172F | 172 | Missense | 4.33 | 2 (1.9%) |
| p.H179R | 179 | Missense | 5.45 | 2 (1.9%) |
| p.I195S | 195 | Missense | 5.35 | 2 (1.9%) |
| p.P222L | 222 | Missense | 2.30 | 2 (1.9%) |
| p.N235S | 235 | Missense | 2.08 | 2 (1.9%) |
| p.R267W | 267 | Missense | 5.97 | 2 (1.9%) |
| p.C277Y | 277 | Missense | 3.06 | 2 (1.9%) |
| p.R283C | 283 | Missense | 1.64 | 2 (1.9%) |
| p.R337C | 337 | Missense | 0.37 | 2 (1.9%) |

**Table 4.** Top 10 most frequent *TP53* variants in *TP53^mono^* with PS ≥ 7.

| **Protein Change** | **Protein Position** | **Variant Classification** | **Phenotypic Score** | **No. of cases** |
| --- | --- | --- | --- | --- |
| p.R175H | 175 | Missense | 12.34 | 12 (11.2%) |
| p.V272M | 272 | Missense | 10.75 | 5 (4.7%) |
| p.V216M | 216 | Missense | 11.13 | 4 (3.7%) |
| p.H193R | 193 | Missense | 11.03 | 3 (2.8%) |
| p.Y220C | 220 | Missense | 8.32 | 3 (2.8%) |
| p.R248W | 248 | Missense | 9.18 | 3 (2.8%) |
| p.C141Y | 141 | Missense | 10.56 | 2 (1.9%) |
| p.Y163C | 163 | Missense | 10.21 | 2 (1.9%) |
| p.C176S | 176 | Missense | 9.93 | 2 (1.9%) |
| p.H179D | 179 | Missense | 16.14 | 2 (1.9%) |
| p.H179Q | 179 | Missense | 21.30 | 2 (1.9%) |
| p.H193L | 193 | Missense | 9.53 | 2 (1.9%) |
| p.H214R | 214 | Missense | 9.80 | 2 (1.9%) |
| p.S215G | 215 | Missense | 7.08 | 2 (1.9%) |
| p.Y220H | 220 | Missense | 7.89 | 2 (1.9%) |
| p.Y234H | 234 | Missense | 11.09 | 2 (1.9%) |
| p.Y234N | 234 | Missense | 10.99 | 2 (1.9%) |
| p.C238Y | 238 | Missense | 11.23 | 2 (1.9%) |
| p.G244D | 244 | Missense | 7.36 | 2 (1.9%) |
| p.G244S | 244 | Missense | 10.70 | 2 (1.9%) |
| p.G245D | 245 | Missense | 9.47 | 2 (1.9%) |
| p.P250L | 250 | Missense | 10.47 | 2 (1.9%) |
| p.R273G | 273 | Missense | 10.11 | 2 (1.9%) |

Most recurrent hotspots mapped close to the DNA-binding interface. Exceptions included *p.Y220* and *p.Y234*, situated outside the immediate DNA-contact region (**Supplementary Figure 9**).

Importantly, functional impact depended not only on codon position but also on the specific amino acid substitution. For example, *p.R248Q* (PS 6.24) and *p.R248W* (PS 9.18) occur at the same codon but fell into different PS groups, mirrored by divergent clinical outcomes (median OS 13.0 vs. 57.2 months), although this difference did not reach statistical significance due to small numbers (HR 2.73, 95% CI 0.60-12.38; p = 0.19; **Supplementary Figure 8B**).

High PS variants cluster almost exclusively within the DBD, with the strongest signals centered around codon 248 (**Figure 2D**).

Additionally, we examined the burden of co-mutations in *TP53* PS-high and PS-low groups. Patients with PS < 7 exhibited a significantly higher number of co-mutations compared with those with PS ≥ 7 (p = 0.001; **Figure 2E, F**), suggesting that *TP53* PS-low cases may more often depend on additional driver alterations.

### Incorporation of VAF further improves the prognostic stratification of monoallelic TP53 variants

Next, we investigated whether incorporating VAF provided additional prognostic information regarding OS. When stratifying monoallelic *TP53* mutations by a 10% VAF threshold, as defined in the 2022 ICC classification [28] to distinguish monoallelic from multi-Hit *TP53* variants, a trend for shorter OS for patients with a high VAF was observed, but statistical significance was not reached (median OS: 16.7 vs 27.2 months, HR = 1.30, 95% CI: 0.90-1.89; p = 0.16; **Supplementary Figure 10A**). The same applied to lower VAF cutoffs (**Supplementary Figure 10B**). Using maximally selected rank statistics [29], the optimal VAF cut-off was determined to be 22% (median OS: 14.3 vs 27.9, HR 1.74, 95% CI: 1.2-2.5; p = 0.002; **Figure 3A**), consistent with previous reports [5]. The same pattern of survival separation was observed when analyses were performed separately within both cohorts. However, statistical significance was narrowly not reached, likely reflecting limited power due to the smaller subset sizes (IWG: HR 1.49, 95% CI: 0.9-2.4; p = 0.1; J-MDS: HR 1.68, 95% CI: 1.0-3.0; p = 0.07, **Supplementary Figure 10C, D**).

**Figure 3.**
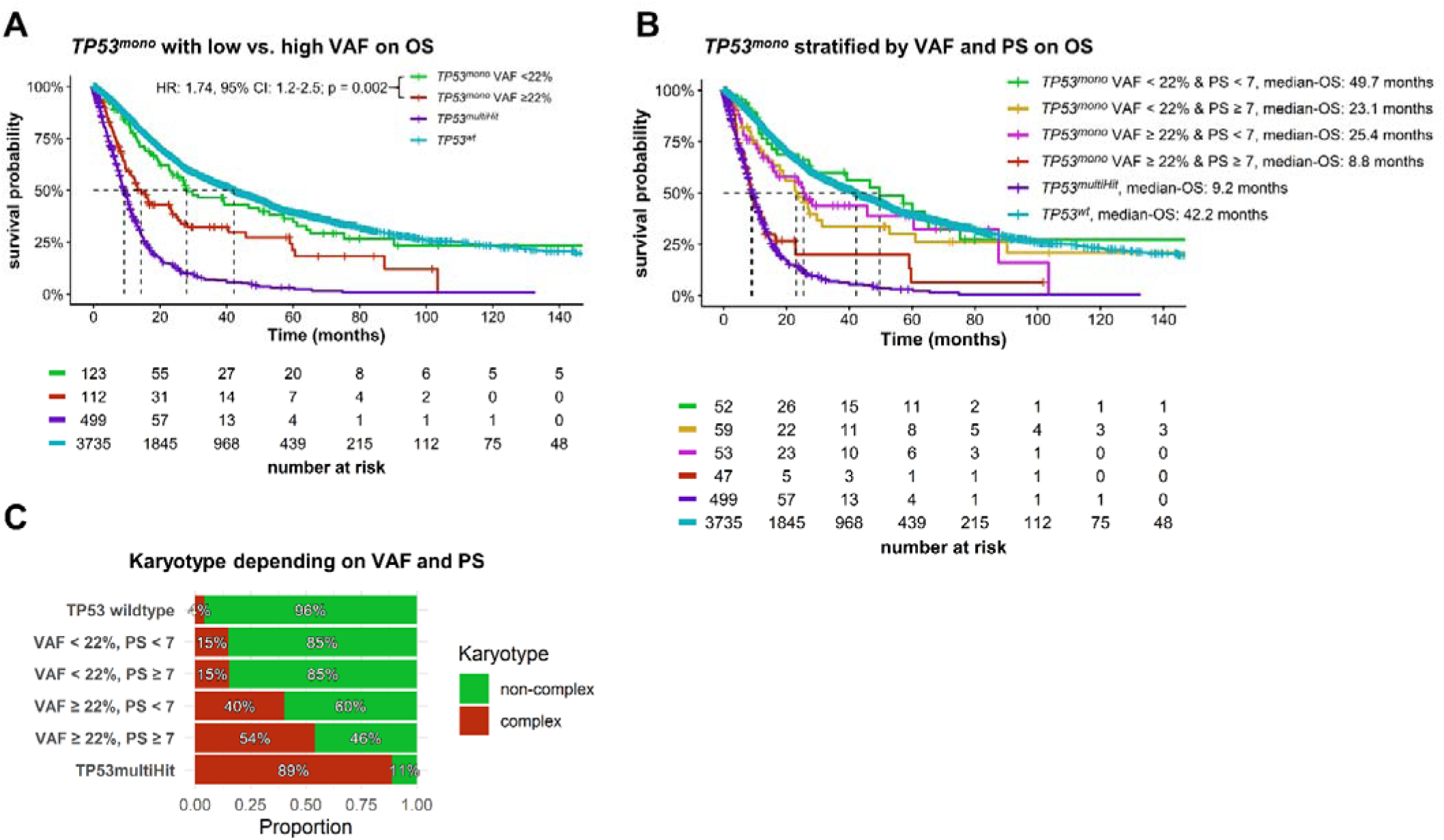
Incorporation of variant allele frequency refines prognostic stratification of *TP53^mono^* mutations. (A) Kaplan-Meier estimates of overall survival (OS) for *TP53^mono^* patients stratified by variant allele frequency (VAF, < 20% vs. ≥20%), shown alongside *TP53^wt^* and *TP53^multiHit^* reference groups. Higher VAF was associated with inferior OS. (B) OS of *TP53^mono^* patients stratified by combined VAF and phenotype score (PS), demonstrating enhanced risk discrimination when both parameters are considered jointly. *TP53^wt^* and *TP53^multiHit^* cases are shown for reference. (C) Proportion of complex versus non-complex karyotypes across *TP53^wt^*, *TP53^mono^* subgroups defined by VAF and PS, and *TP53^multiHit^* cases, illustrating an increasing frequency of complex karyotypes with higher VAF and PS.

Importantly, a linear regression analysis showed no correlation between PS and VAF, confirming that the prognostic contributions of both factors are independent of one another (R² = 0.01; **Supplementary Figure 10F**).

Further, we evaluated whether combining VAF and PS could enhance prognostic discrimination. Applying a VAF cutoff of 22% together with a PS cutoff of 7 resulted in a strong separation of survival curves (VAF <22% & PS <7 vs. VAF ≥22% & PS ≥7: HR 3.3, 95% CI: 1.9-5.7, p < 0.001, **Figure 3B**). Patients with VAF ≥22% and PS ≥7 had an OS comparable to that of MDS patients with *TP53^multiHit^* mutations (median OS: 8.8 vs. 9.2 months, p = 0.2), whereas patients with VAF <22% and PS <7 exhibited an OS similar to *TP53^wt^* cases (median OS: 49.7 vs. 42.2 months, p = 0.9, **Figure 3B**). This strong prognostic value was observed in both cohorts (VAF <22% & PS <7 vs. VAF ≥22% & PS ≥7 for IWG: HR 2.84, 95% CI: 1.2-.6.6, p = 0.01; J-MDS: HR 2.46, 95% CI: 1.0-6.1, p = 0.05, **Supplementary Figure 11**).

Analysis of complex karyotype frequency across these four subgroups revealed marked differences. In contrast to OS, VAF appeared to have a stronger impact on the occurrence of complex karyotypes (**Figure 3C**).

### Prognostic impact of mutation-type combinations and VAF in TP53^multiHit^ disease

We then investigated whether the type of *TP53^multiHit^*mutation combination affected OS. No significant differences in OS were observed across *TP53^multiHit^* subgroups defined by mutation type combinations, deletion + missense (del+ms, median OS: 8.7 months), frameshift + missense (median OS: 10.1 months), missense + missense (ms+ms, median OS: 10.3 months), nonsense + missense (median OS: 6.3 months), or splice site + missense (10.0 months) (log-rank overall p = 0.3, **Figure 4A**).

**Figure 4.**
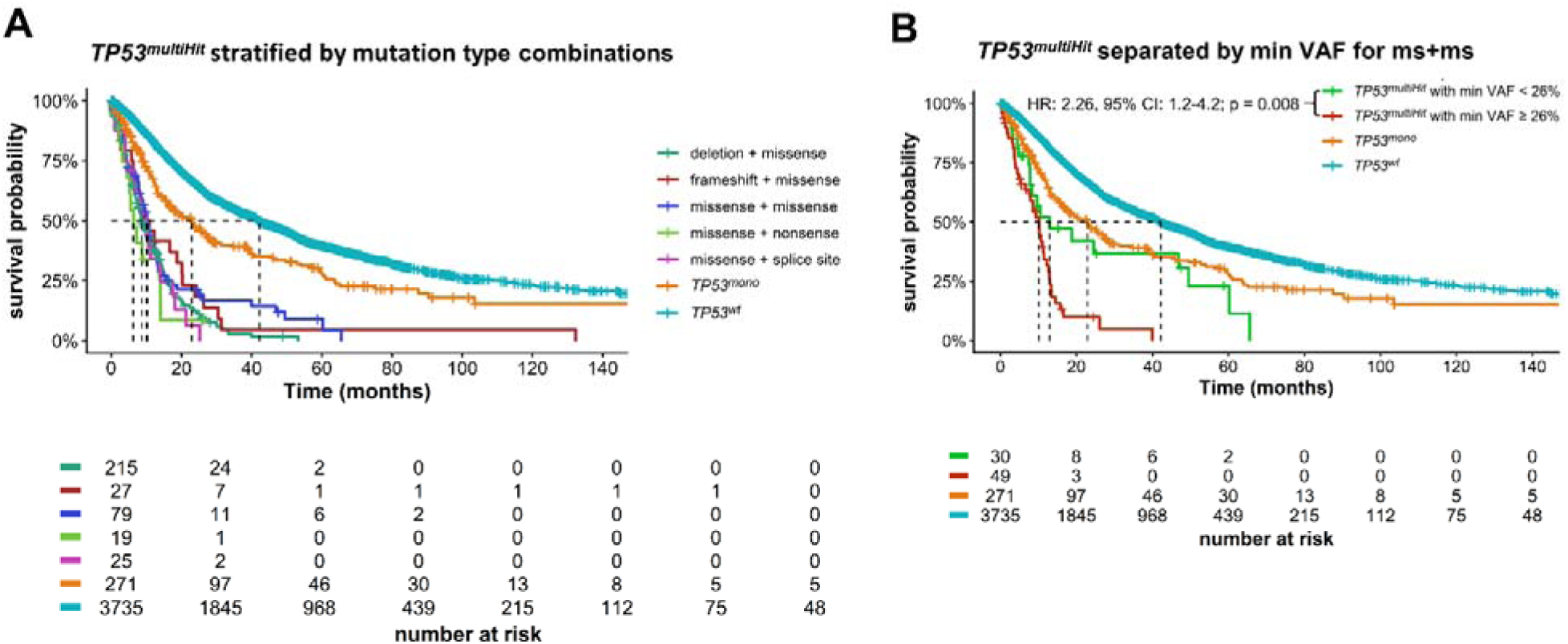
Mutation type and clonal burden do not uniformly modify prognosis in *TP53^multiHit^* disease. (A) Kaplan-Meier estimates of overall survival (OS) in *TP53^multiHit^*patients stratified by mutation type combinations, including missense + missense, frameshift + missense, nonsense + missense, splice-site + missense, and deletion + missense configurations. *TP53^mono^* and *TP53^wt^*cases are shown for reference. No significant differences in OS were observed across mutation type combinations. (B) OS of *TP53^multiHit^*patients stratified by the lower VAF, demonstrating significant survival separation at a variant allele frequency (VAF) cutoff of 26%. *TP53^mono^*and *TP53^wt^* cases are shown for reference.

Next, we focused on ms+ms cases to assess whether the type of missense mutation influenced outcomes according to the PS. Using a PS cutoff of 7, patients were stratified based on the higher, average, or lower PS value of the two mutations. None of these approaches resulted in a significant difference in OS, exemplified for the lower PS value with a cutoff of 7 (**Supplementary Figure 12A**). Notably, when stratifying by the lower VAF, a significant prognostic prediction was observed for VAF thresholds between 22% and 29%, with the strongest effect seen at 26% (HR 2.26, 95% CI: 1.23-4.18; p = 0.008, **Figure 4B**). The same analysis was performed for del+ms combinations. Stratification by VAF revealed the strongest numerical separation of survival curves at a cutoff of 60% using maximally selected rank statistics [29] (HR 1.25, 95% CI: 0.9-1.7; p = 0.2, **Supplementary Figure 12B**).

### Phenotype Score is an independent prognostic factor

To further characterize prognostic determinants in *TP53^mono^* disease, we performed univariable and multivariable Cox proportional hazards regression analyses of established clinical and genetic risk factors, including sex, age, peripheral blood counts, bone marrow blast percentage, complex karyotype, and IPSS-R and IPSS-M, for OS.

In univariable analyses, both a high *TP53* PS (PS ≥ 7; HR 1.75, p = 0.003) and higher *TP53* VAF (VAF ≥ 22%; HR 1.74, p = 0.002) were significantly associated with inferior OS (**Table 5**). These variables, together with other significant clinical covariates, were subsequently entered into multivariable models. To avoid multicollinearity, composite prognostic scores (IPSS-R or IPSS-M) were not combined with their individual components in multivariable analyses.

**Table 5.** Univariable Analysis.

| **Variable [unit]** | **HR (95% CI)** | **p-value** |
| --- | --- | --- |
| PS ($\geq 7$) | 1.75 (1.21-2.53) | 0.003 |
| VAf ($\geq 22\%$) | 1.74 (1.23-2.46) | 0.002 |
| Age [years] | 1.02 (1.01-1.04) | 0.004 |
| Sex (male) | 1.44 (1.04-2.01) | 0.03 |
| Hb [g/dL] | 0.92 (0.85-1) | 0.05 |
| WBC [$\times 10^9/L$] | 1.03 (1.02-1.05) | < 0.001 |
| ANC [$\times 10^9/L$] | 1.04 (0.99-1.09) | 0.15 |
| Plt [$\times 10^9/L$] | 0.997 (0.996-0.999) | < 0.001 |
| Bone marrow blasts [%] | 1.02 (1.01-1.04) | 0.003 |
| Complex karyotype | 2.29 (1.63-3.23) | < 0.001 |
| IPSS-R category | 1.52 (1.32-1.74) | < 0.001 |
| IPSS-M category | 1.29 (1.16-1.43) | < 0.001 |

In this model, the *TP53* PS remained independently associated with OS (HR 1.60, p = 0.02), whereas *TP53* VAF ≥22% no longer retained statistical significance, particularly after adjustment for complex karyotype (HR 1.27, p = 0.27, **Table 6**).

**Table 6.** Multivariable Analysis Variable [unit].

| **Variable [unit]** | **HR** | **p-value** |
| --- | --- | --- |
| PS ($\geq 7$) | 1.60 (1.08-2.78) | 0.02 |
| VAf ($\geq 22\%$) | 1.27 (0.83-1.95) | 0.27 |
| Age [years] | 1.02 (1-1.04) | 0.03 |
| Sex (male) | 1.38 (0.90-2.11) | 0.14 |
| WBC [$\times 10^9/L$] | 1.05 (1.02-1.09) | 0.004 |
| Plt [$\times 10^9/L$] | 0.998 (0.997-1.000) | 0.05 |
| Complex karyotype | 2.31 (1.44-3.71) | < 0.001 |

Collectively, these findings indicate that the *TP53* PS captures prognostic information beyond established clinical and cytogenetic risk factors and represents an independent determinant of survival in *TP53^mono^* MDS, whereas *TP53* VAF alone does not confer independent prognostic value after adjustment.

## Discussion

In this study from two large independent cohorts, we confirmed that *TP53* allelic state is a major determinant of outcome, with *TP53^multiHit^* conferring the poorest survival and *TP53^mono^* showing intermediate prognosis. Notably, *TP53^mono^*mutations exhibit marked biological heterogeneity. Incorporating functional variant annotation via the PS identified a clinically relevant subgroup of *TP53^mono^*-mutated patients with markedly inferior outcomes.

While *TP53^mono^* alterations are often treated as a single risk category, our data demonstrate that monoallelic patients carrying high PS variants (PS ≥7) showed pronounced shorter OS than those with low PS, despite sharing the same allelic state. This supports a model in which some *TP53^mono^* variants, particularly those with strong dominant-negative or severe loss-of-function behavior, can phenocopy a more aggressive biology even before acquisition of a second hit. Especially within lower-risk disease, a high PS was associated with substantial survival differences. These findings suggest that these patients may warrant closer monitoring, earlier therapeutic escalation, or prioritization for transplant evaluation in selected patients.

We observed that *TP53^mono^* lesions were predominantly missense mutations clustered within DBD, consistent with prior reports [17]. Our data extend these observations by linking DBD clustering to quantitative functional impairment and clinical outcome.

An intriguing variant-level observation is the enrichment of *TP53* alterations affecting codon *p.Y220* in the multi-hit setting, whereas *p.Y220* variants were comparatively uncommon among monoallelic cases.

The preferential representation of *p.Y220* within multi-hit configurations may indicate that certain *TP53* variants confer maximal clonal advantage only in the context of additional *TP53* hits. These data support the concept that *TP53* evolution in MDS is not only allelic state-dependent but also variant-dependent, and that specific hotspots may reflect distinct evolutionary trajectories. Notably, PS-high cases were not characterized by a higher overall mutational burden. This supports a model in which adverse biology in this subgroup is primarily driven by the *TP53* lesion itself, rather than by the accumulation of additional disease-driving mutations.

We further demonstrate that PS and VAF capture non-redundant biological dimensions. The absence of correlation between PS and VAF indicates that functional severity and clonal burden are distinct. While combining PS with higher VAF thresholds improved risk stratification and identified subgroups with outcomes approximating *TP53^wt^* or *TP53^multiHit^*disease, multivariable analysis showed that PS retained independent prognostic significance whereas VAF did not. Consistently, PS showed no clear association with complex karyotype, whereas higher VAF was strongly enriched in cases with complex karyotype. Thus, PS primarily reflects functional disruption of *TP53*, whereas VAF largely captures clonal dominance and disease evolution.

Importantly, we performed an in-depth comparative analysis between the two large, independent cohorts to better understand the survival differences observed among *TP53^mono^*-mutated patients. Systematic analysis of *TP53* allelic state, variant composition, phenotypic severity, and clonal burden showed that the Japanese cohort had overall higher *TP53* PS and VAF levels and a higher proportion of both *TP53^mono^* and *TP53^multiHit^*cases. These differences were not driven by enrichment of specific *TP53* hotspot variants, but rather reflected a broader shift in the functional and clonal characteristics of *TP53* alterations, potentially indicating differences in underlying disease biology between cohorts while supporting the robustness of PS-based stratification across clinically heterogeneous populations.

Several limitations should be considered. First, PS values were derived from high-throughput functional screens performed in a defined experimental system. While these assays provide systematic functional resolution across *TP53* variants, context-dependent effects such as cell type, cellular stress conditions and interactions with the immune microenvironment may not be fully captured. Second, PS values were generated in a single cellular model and may not fully reflect *TP53* function in hematopoietic stem and progenitor cells. Finally, although the overall cohort size was large, statistical power was limited for rare *TP53* variants and certain subgroup analyses.

Overall, our findings support a refined view of *TP53*-mutated MDS in which allelic state remains critical, but variant-level functional heterogeneity within monoallelic disease is clinically relevant. Incorporation of functional mutation metrics such as the PS and VAF into clinical annotation may enhance existing risk frameworks and enable more individualized prognostication and treatment planning.

## Data Availability

Data from the International Working Group (IWG) cohort is available as described in prior publications. Deidentified individual participant data from the Japanese J-MDS cohort are available upon reasonable request and subject to institutional approval and data transfer agreements.

## Acknowledgments

We acknowledge the International Working Group (IWG) for Myelodysplastic Neoplasms and participating centers for providing access to clinical and molecular data. We further thank the contributing institutions of the Japanese MDS cohort for data generation and curation.

A.S. is a clinician scientist of the ICON program at the medical faculty Mannheim of Heidelberg University, DN is funded by the „Deutsche Forschungsgemeinschaft (DFG), CRC1709 and was an endowed Professor of the German José-Carreras-Foundation (DJCLSH03/01). N.S. is supported by the Medical Faculty Mannheim of the Heidelberg University “SEED” and the Torsten Haferlach Leukemia Diagnostics Foundation. V.R. is funded by the Health + Life Science Alliance Heidelberg Mannheim and received state funds approved by the State Parliament of Baden-Württemberg.

This work was supported by grants from the Japan Agency for Medical Research and Development (AMED): nos. JP24tk0124003 to S.O, JP24ck0106791, JP25ck0106019, and JP25kk0305028 to Y.O., JP22ck0106691 and JP24ck0106875 to Y.O., Y.N., and S.O; the Moonshot Research and Development Program (no. JP24zf0127009) to S.O.; the Japan Society for the Promotion of Science (JSPS): KAKENHI (nos. JP24H00009 to S.O, JP24K19223 to Y.O.); the Japan Science and Technology Agency (JST): Fusion Oriented Research for disruptive Science and Technology (FOREST) Program (no. JPMJFR220L) to Y.O.

## Authorship Contributions

A.S., D.N., N.S. and V.R. conceived and designed the study. A.S. performed the analyses and drafted the manuscript. Y.O., Y.N., and S.O. contributed patient data and provided critical input on study design and data interpretation. E.H. contributed to figure preparation and data visualization. W.-K.H. provided infrastructure and institutional support. L.S., M.A., G.M., E.A., F.R., and V.N. contributed to data acquisition and critically revised the manuscript. All authors reviewed, edited, and approved the final version of the manuscript.

---

**Supplementary Figure 1.**
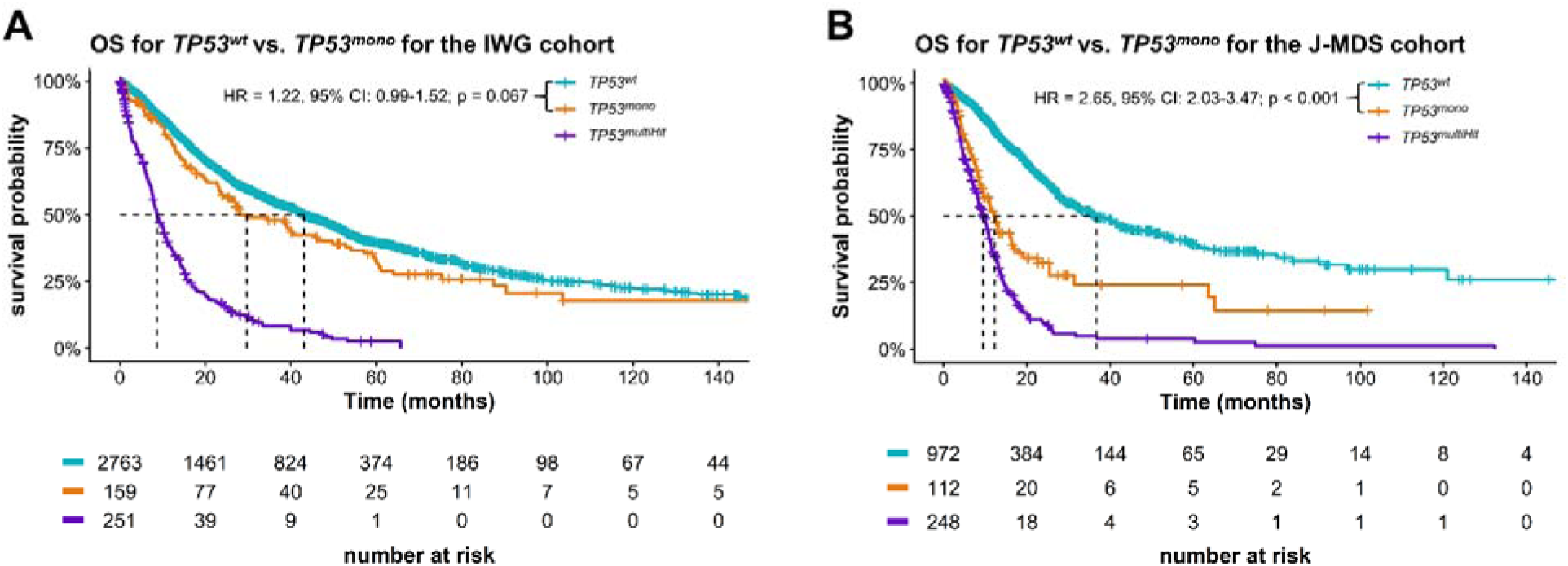
Overall survival according to *TP53* allelic status in individual cohorts. (A) Kaplan-Meier estimates of overall survival (OS) for *TP53^wt^*, *TP53^mono^*, and *TP53^multiHit^* patients in the IWG cohort. *TP53^wt^* and *TP53^mono^* patients showed similar survival outcomes. (B) Corresponding OS analysis in the independent J-MDS cohort, demonstrating a significant survival difference between *TP53^wt^*and *TP53^mono^* cases.

**Supplementary Figure 2.**
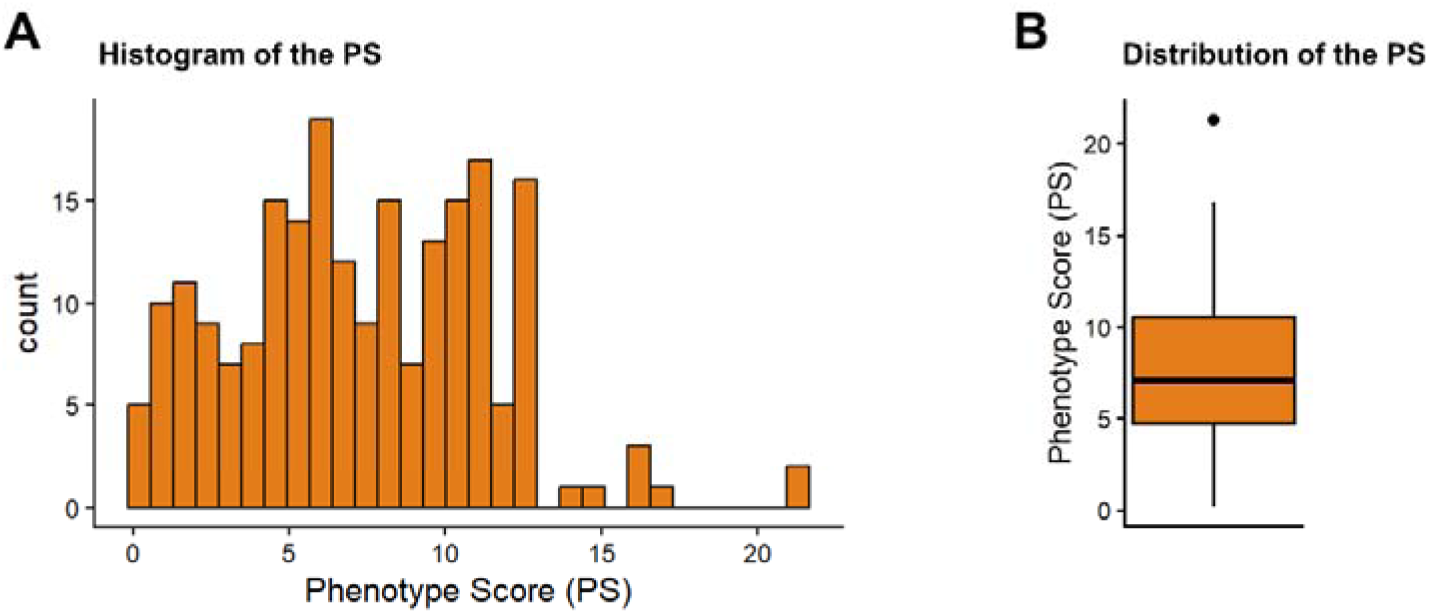
Distribution of the phenotype score in *TP53^mono^* cases. (A) Histogram showing the distribution of phenotype score (PS) values. (B) Boxplot summarizing the distribution of PS values.

**Supplementary Figure 3.**
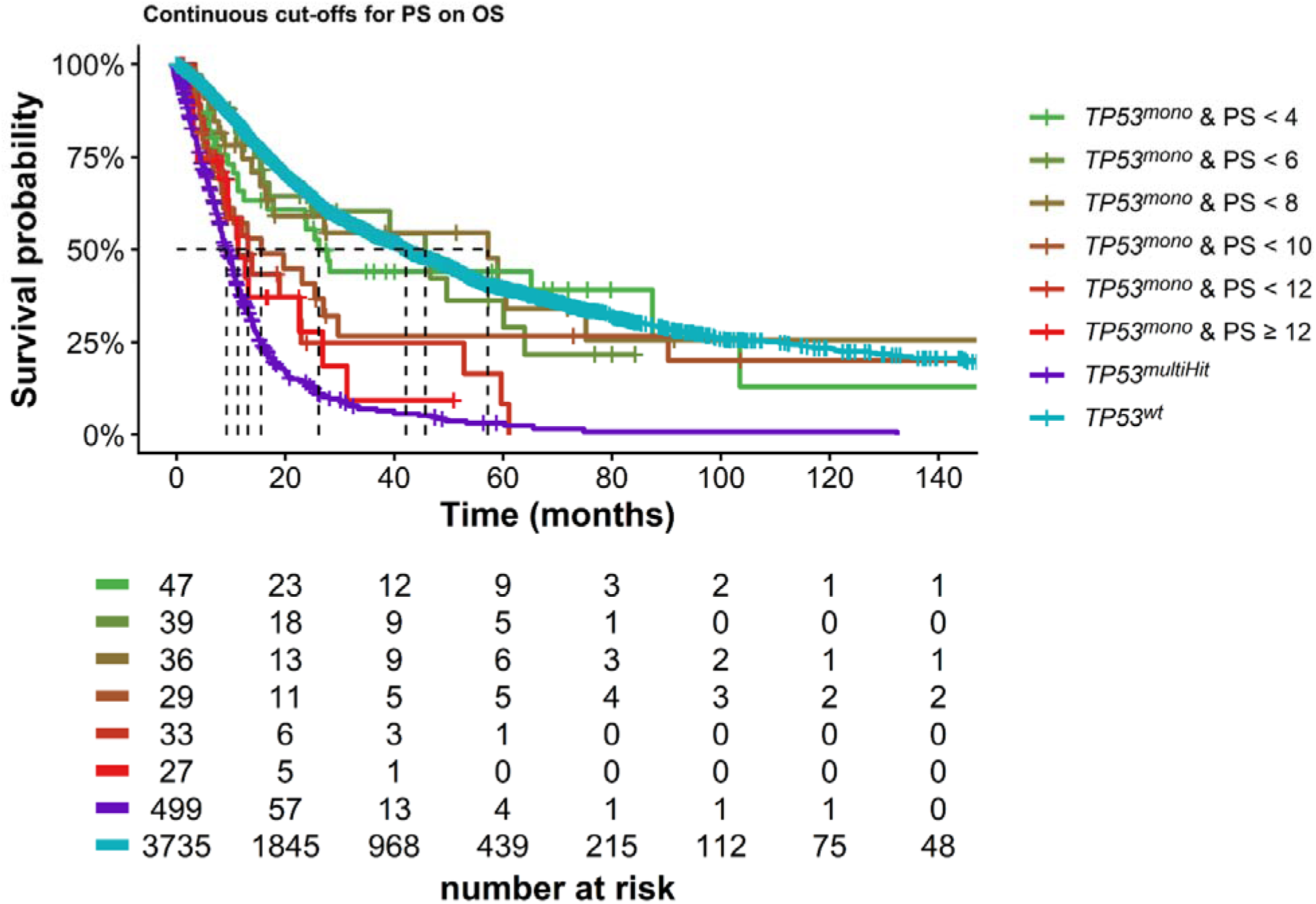
Continuous phenotype score cut-offs and their association with overall survival in *TP53^mono^* cases. Kaplan-Meier estimates of overall survival (OS) for *TP53^mono^* patients stratified by increasing phenotype score (PS) cut-offs (<4, <6, <8, <10, <12 and ≥12). *TP53^wt^* and *TP53^multiHit^*cases are shown for reference.

**Supplementary Figure 4.**
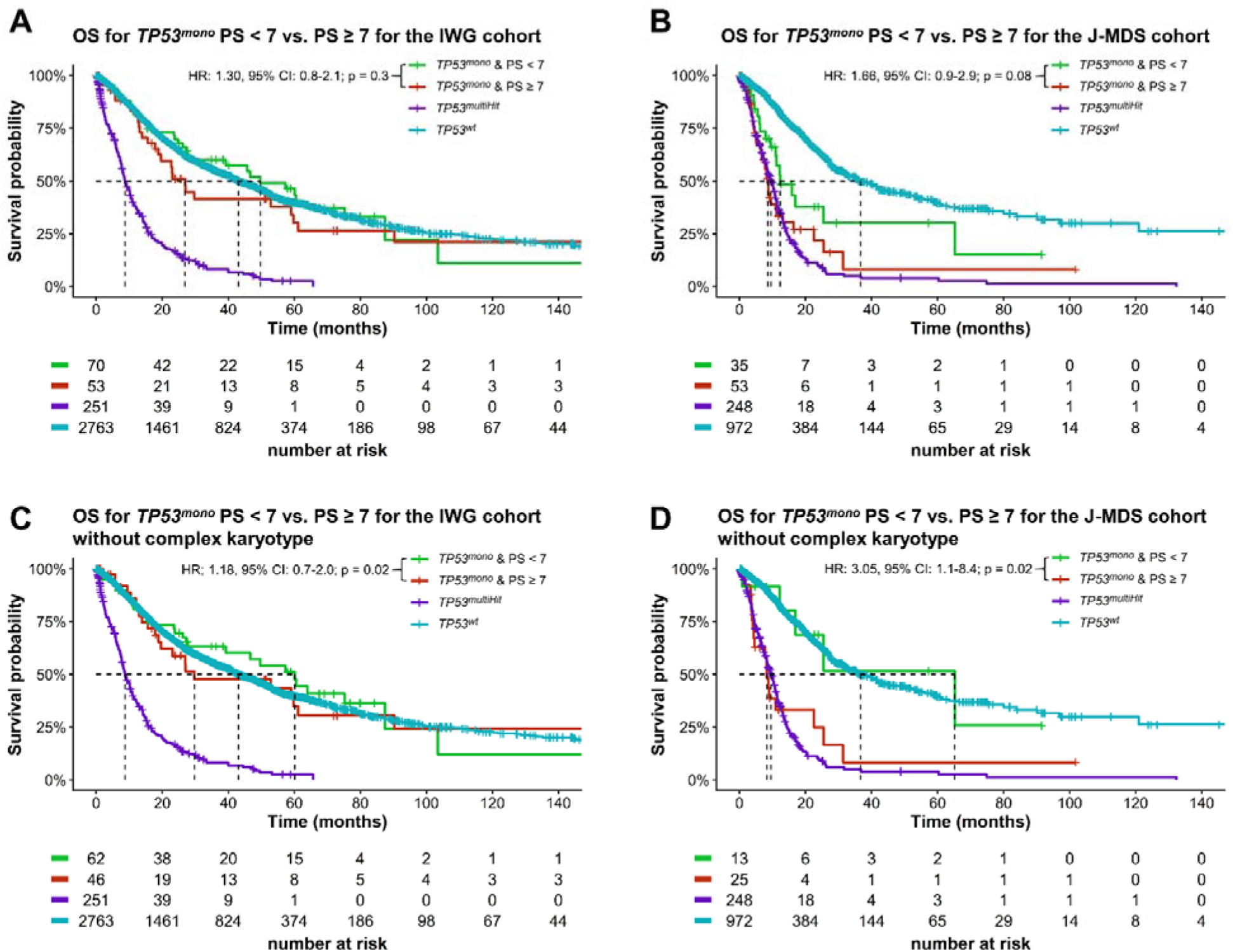
Overall survival of *TP53^mono^*cases stratified by phenotype score within individual cohorts and after exclusion of complex karyotype. (A) Overall survival (OS) for *TP53^mono^* patients with PS < 7 vs. PS ≥ 7 in the IWG cohort. (B) Corresponding OS analysis in the J-MDS cohort. (C) OS for *TP53^mono^*patients with phenotype score (PS) < 7 vs. PS ≥ 7 in the IWG cohort after exclusion of cases with complex karyotype. (D) Corresponding analysis in the J-MDS cohort after exclusion of complex karyotype.

**Supplementary Figure 5.**
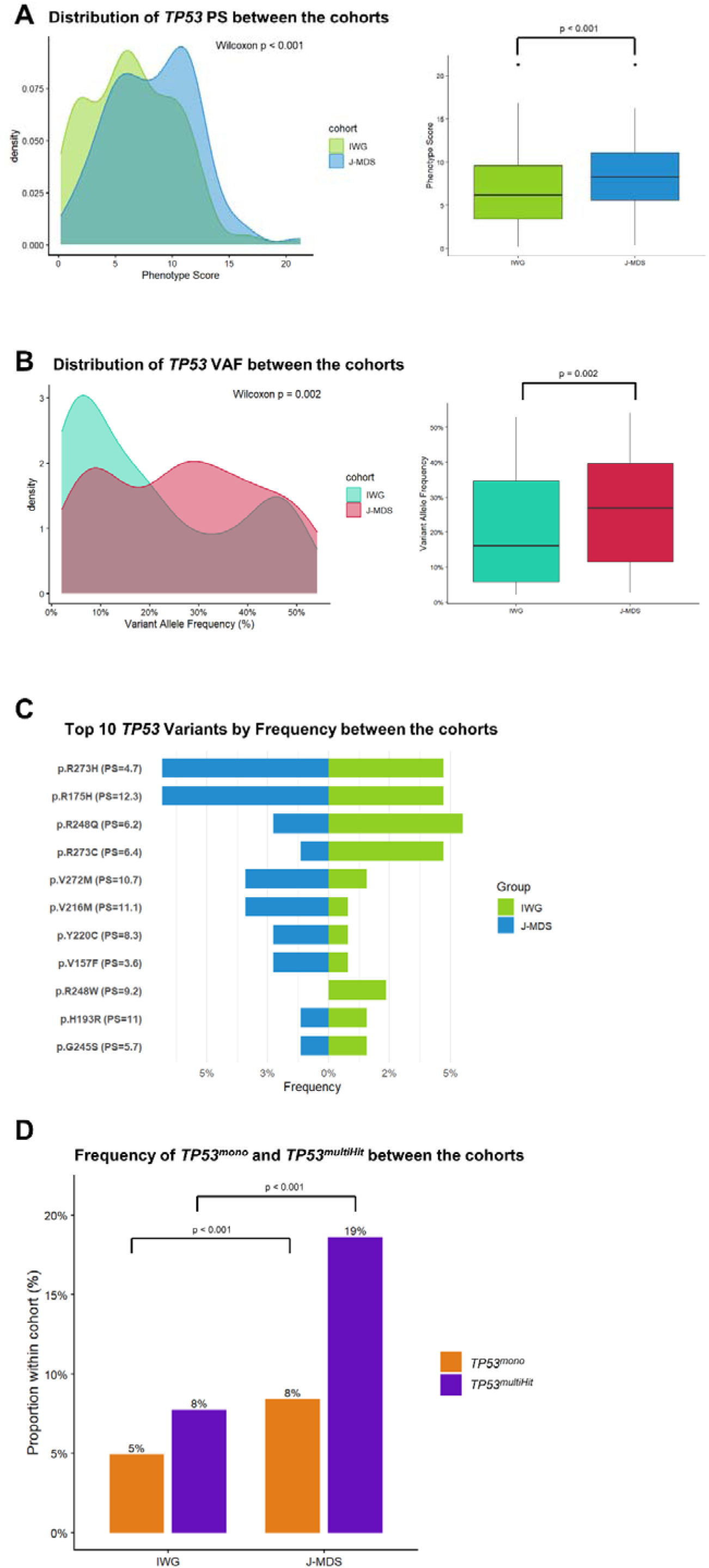
Comparison of *TP53* phenotype score, variant allele frequency and mutation frequencies between the IWG and J-MDS cohorts. (A) Distribution of *TP53* PS values between cohorts, showing significantly higher phenotype score (PS) values in the J-MDS cohort (p < 0.001). (B) Distribution of *TP53* VAF between cohorts, demonstrating higher VAF levels in the J-MDS cohort (p = 0.002). (C) Top ten most frequent *TP53* variants stratified by cohort, showing no significant enrichment of individual variants between cohorts. (D) Proportion of *TP53^mono^*and *TP53^multiHit^* cases within each cohort, illustrating higher frequencies of both *TP53^mono^* and *TP53^multiHit^* in the J-MDS cohort.

**Supplementary Figure 6.**
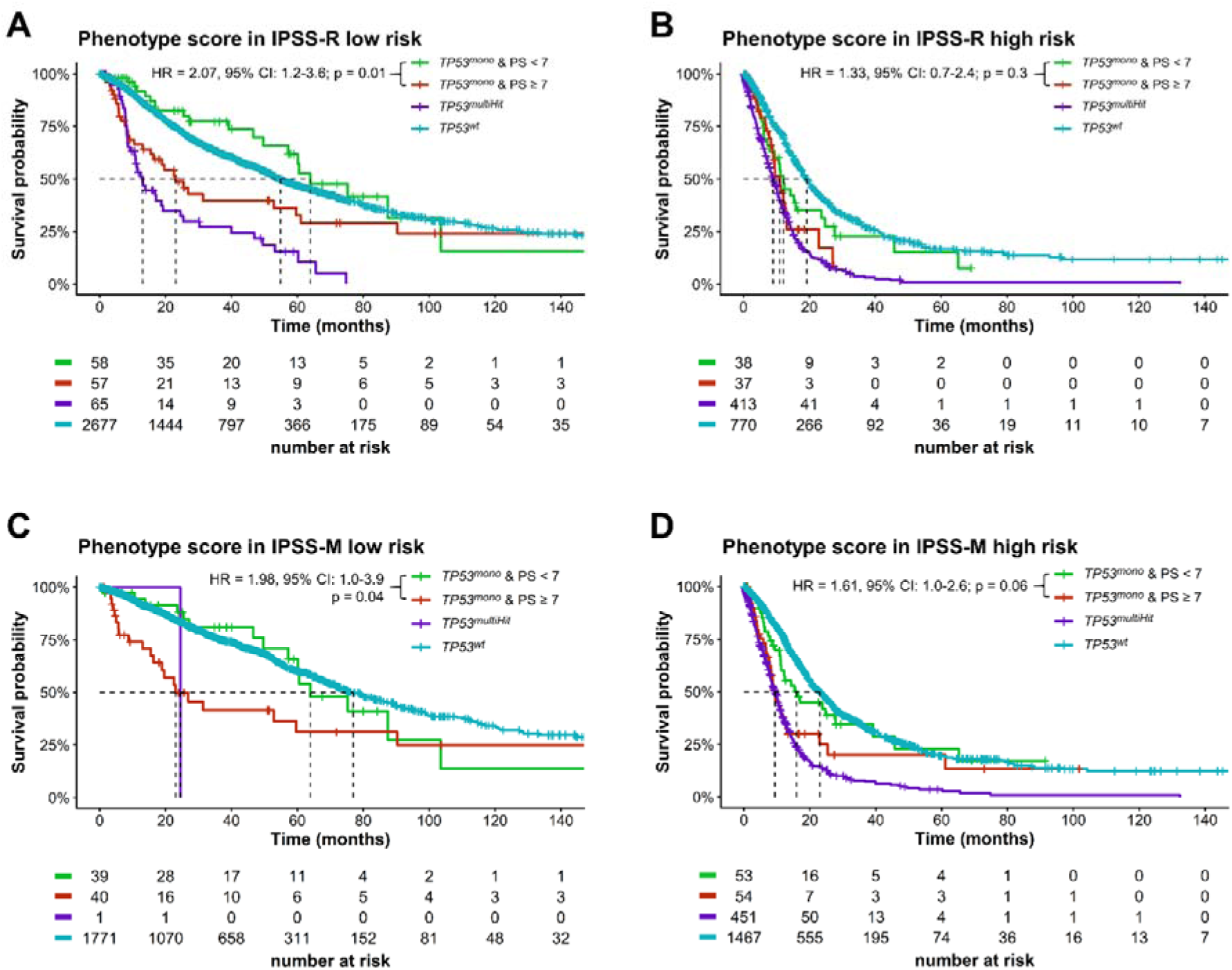
Overall survival according to phenotype score within IPSS-R and IPSS-M risk categories. (A, C) In lower-risk IPSS-R and IPSS-M groups, phenotype score (PS) ≥7 identifies patients with inferior overall survival (OS). (B, D) In higher-risk groups, PS-based separation is less pronounced.

**Supplementary Figure 7.**
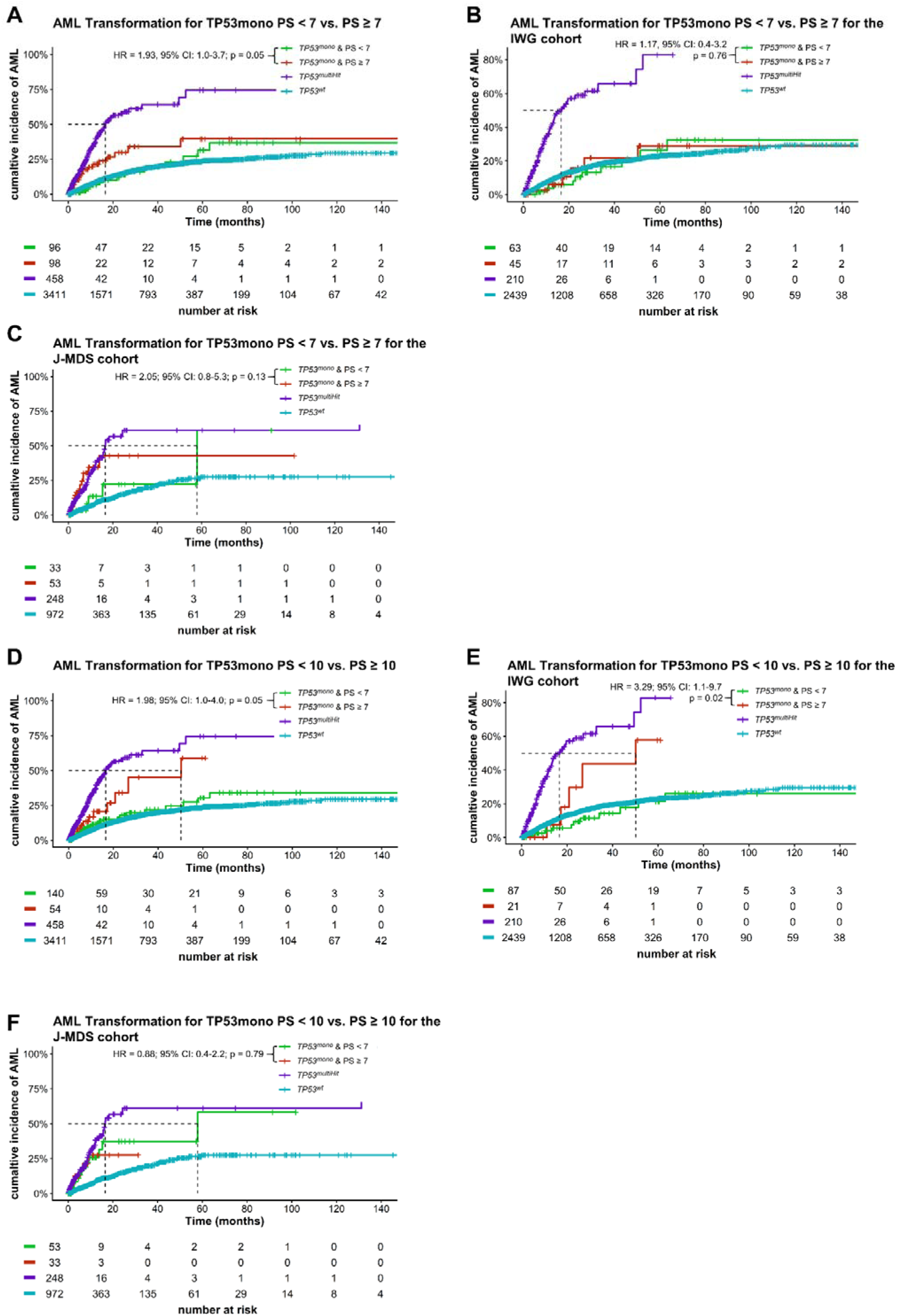
Cumulative incidence of acute myeloid leukemia according stratified by phenotype score in *TP53^mono^* cases. (A) *TP53^mono^* patients with phenotype score (PS) ≥7 show a higher cumulative incidence of AML compared with PS <7. (B-C) Cohort-specific analyses in the IWG and J-MDS cohorts demonstrate similar trends. (D-F) A higher PS threshold (≥10) further stratifies acute myeloid leukemia (AML) risk, particularly in the IWG cohort.

**Supplementary Figure 8.**
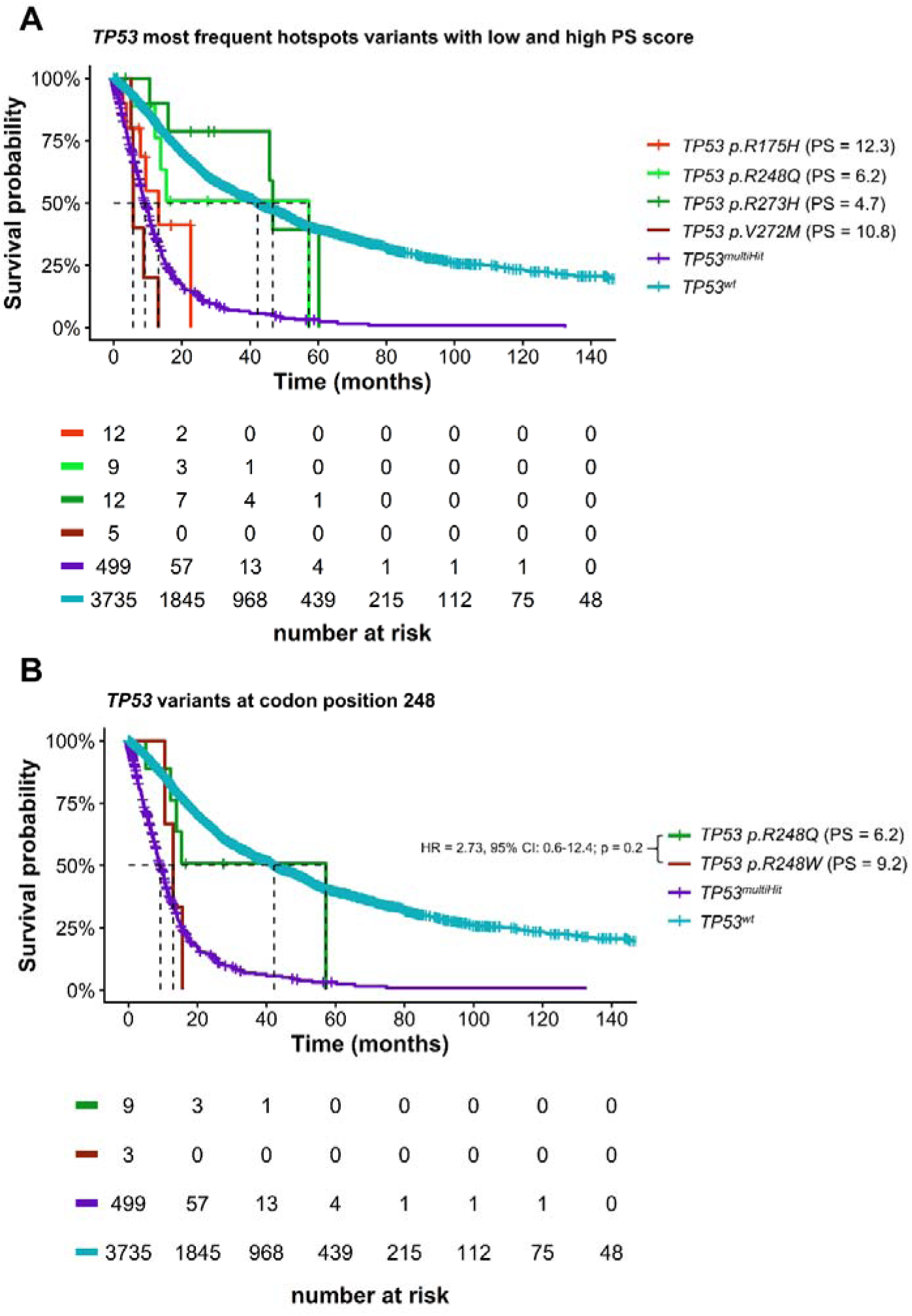
Survival according to recurrent *TP53* hotspot variants and codon-specific substitutions. (A) Overall survival (OS) of *TP53^mono^* patients harboring the most frequent hotspot variants stratified by phenotype score (PS) group. (B) OS of *TP53^mono^* patients with variants affecting codon 248 (*p.R248Q* vs. *p.R248W*).

**Supplementary Figure 9.**
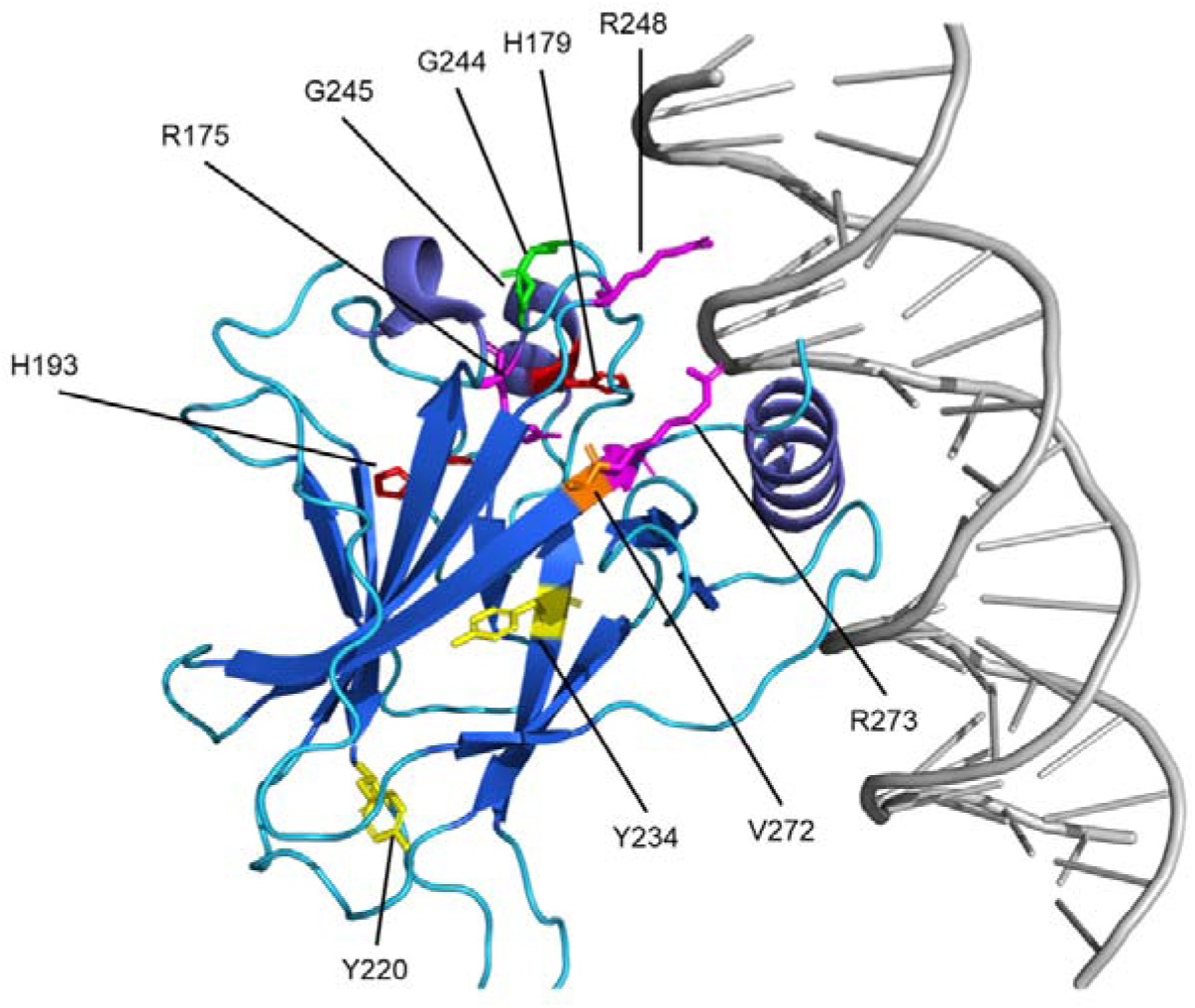
Structural localization of recurrent *TP53* variants. Three-dimensional structure of *TP53* bound to DNA, illustrating the spatial distribution of recurrent hotspot variants and their proximity to the DNA-binding interface, except for p.Y220 and p.Y234. Residues are color-coded by amino acid: glycine (green), arginine (magenta), histidine (red), tyrosine (yellow), and valine (orange).

**Supplementary Figure 10.**
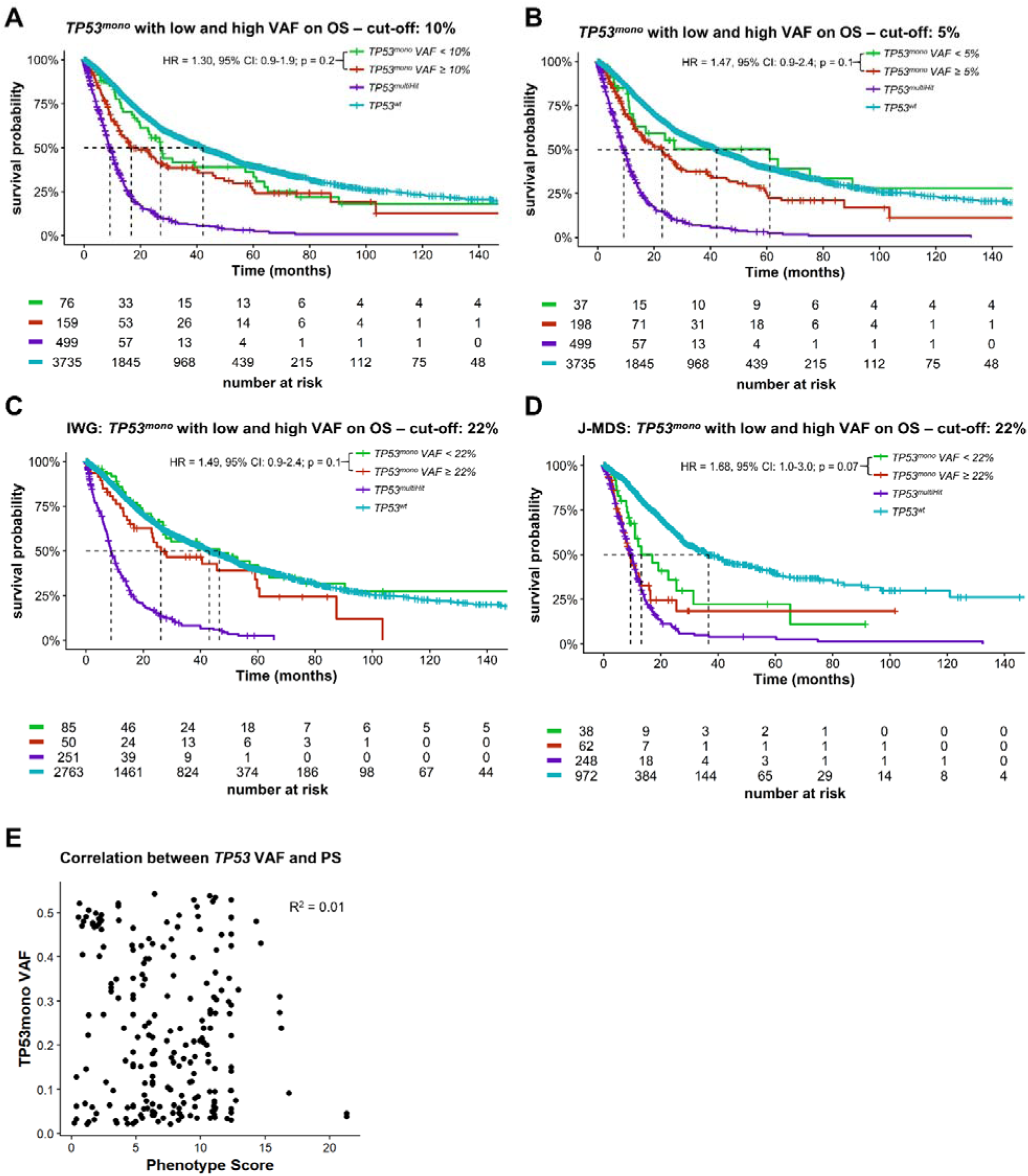
Overall survival according to variant allele frequency (VAF) thresholds and correlation between VAF and phenotype score in *TP53^mono^*cases. (A) Overall survival (OS) stratified by variant allele frequency (VAF) <10% vs. VAF ≥10%, showing no statistically significant separation. (B) OS stratified by VAF <5% vs. VAF ≥5%, showing no statistically significant separation. (C) OS stratified by VAF <22% vs. VAF ≥22% in the overall cohort, showing a significant survival difference. (D) Corresponding analysis in the IWG cohort (cut-off 22%), showing a trend toward inferior OS with higher VAF that did not reach statistical significance. (E) Corresponding analysis in the J-MDS cohort (cut-off 22%), showing a similar trend that did not reach statistical significance. (F) Scatter plot of *TP53* VAF versus phenotype score (PS), showing no meaningful correlation.

**Supplementary Figure 11.**
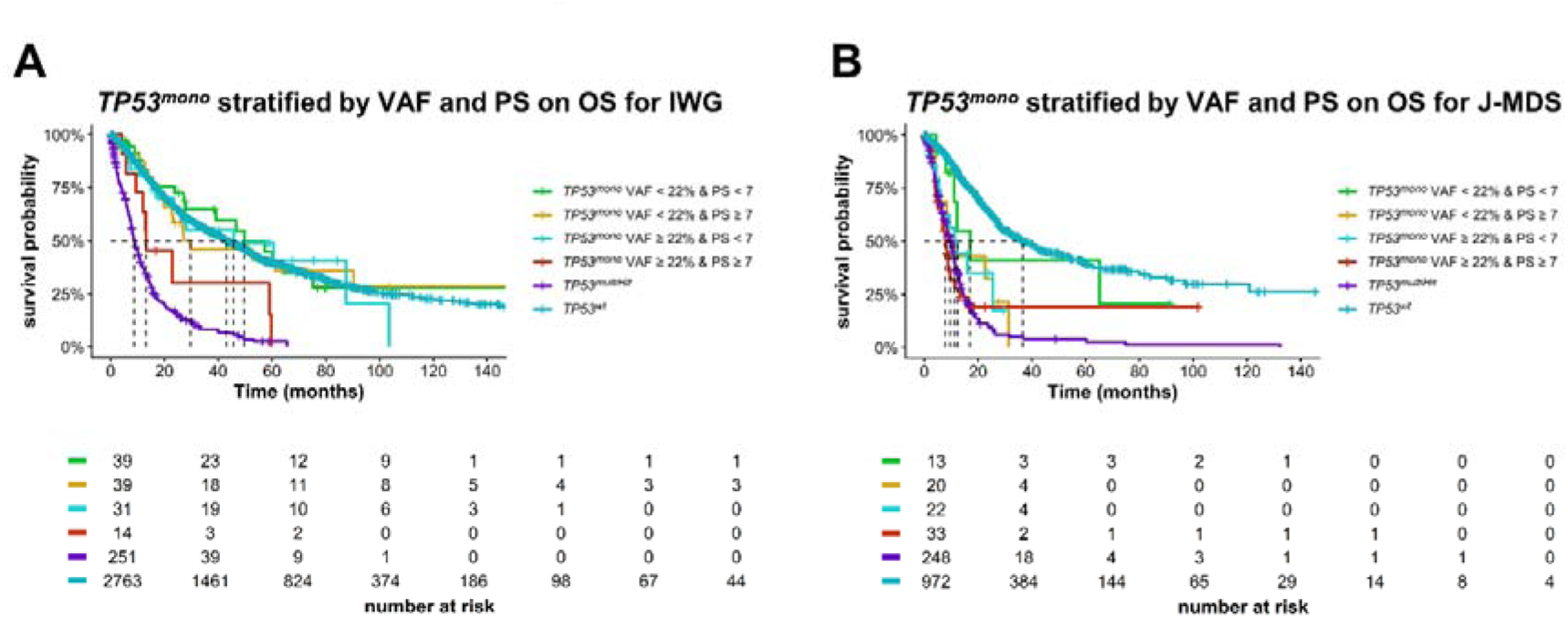
Overall survival according to combined variant allele frequency and phenotype score stratification in *TP53^mono^*cases within individual cohorts. (A) *TP53^mono^*patients in the IWG cohort stratified by variant allele frequency (VAF, <22% vs. ≥22%) and phenotype score (PS, <7 vs. ≥7), demonstrating inferior overall survival (OS) in patients with both high VAF and high PS. (B) Corresponding analysis in the J-MDS cohort, showing a similar pattern of risk stratification based on combined VAF and PS.

**Supplementary Figure 12.**
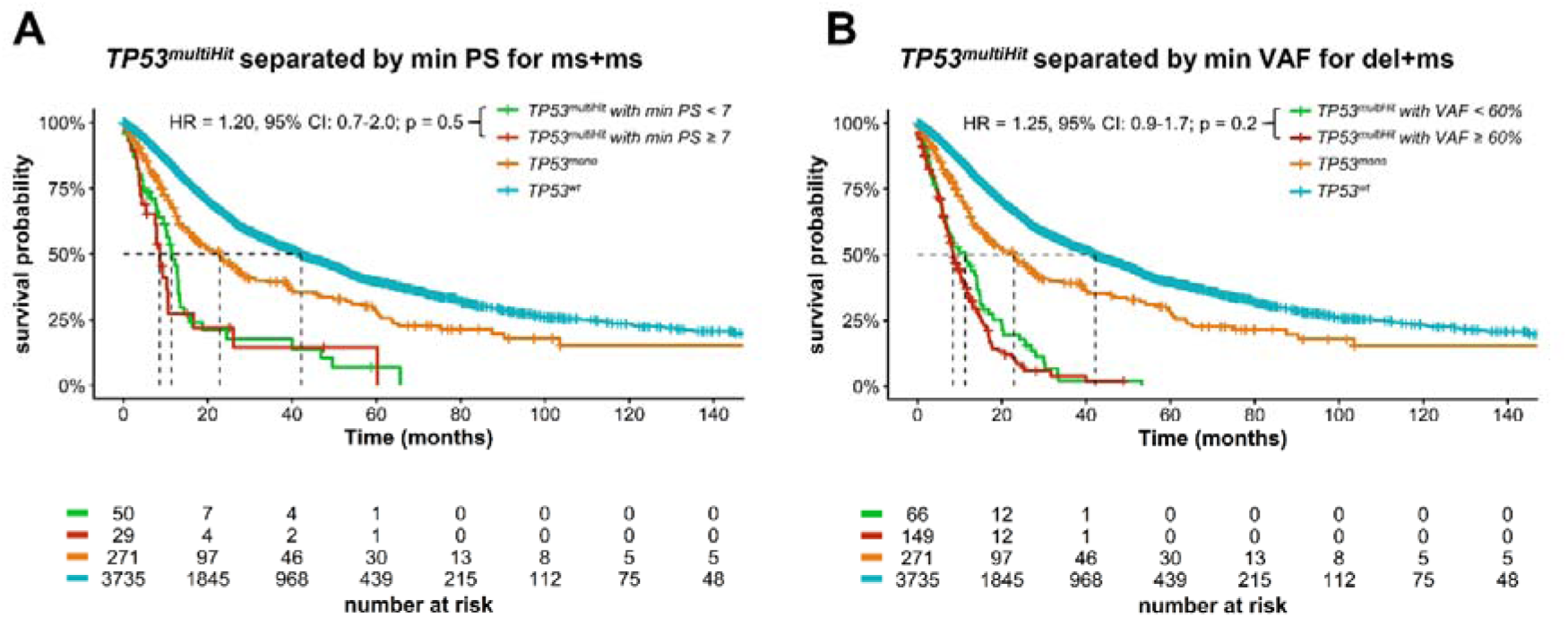
Overall survival in *TP53^multiHit^* cases according to mutation type–specific phenotype score and variant allele frequency stratification. (A) *TP53^multiHit^* cases with missense + missense (ms+ms) combinations stratified by the lower phenotype score (PS, <7 vs. ≥7), showing no significant difference in overall survival (OS). (B) *TP53^multiHit^* cases with deletion + missense (del+ms) combinations stratified by the lower VAF (<60% vs. ≥60%), showing no statistically significant survival separation.

**Table S1.**
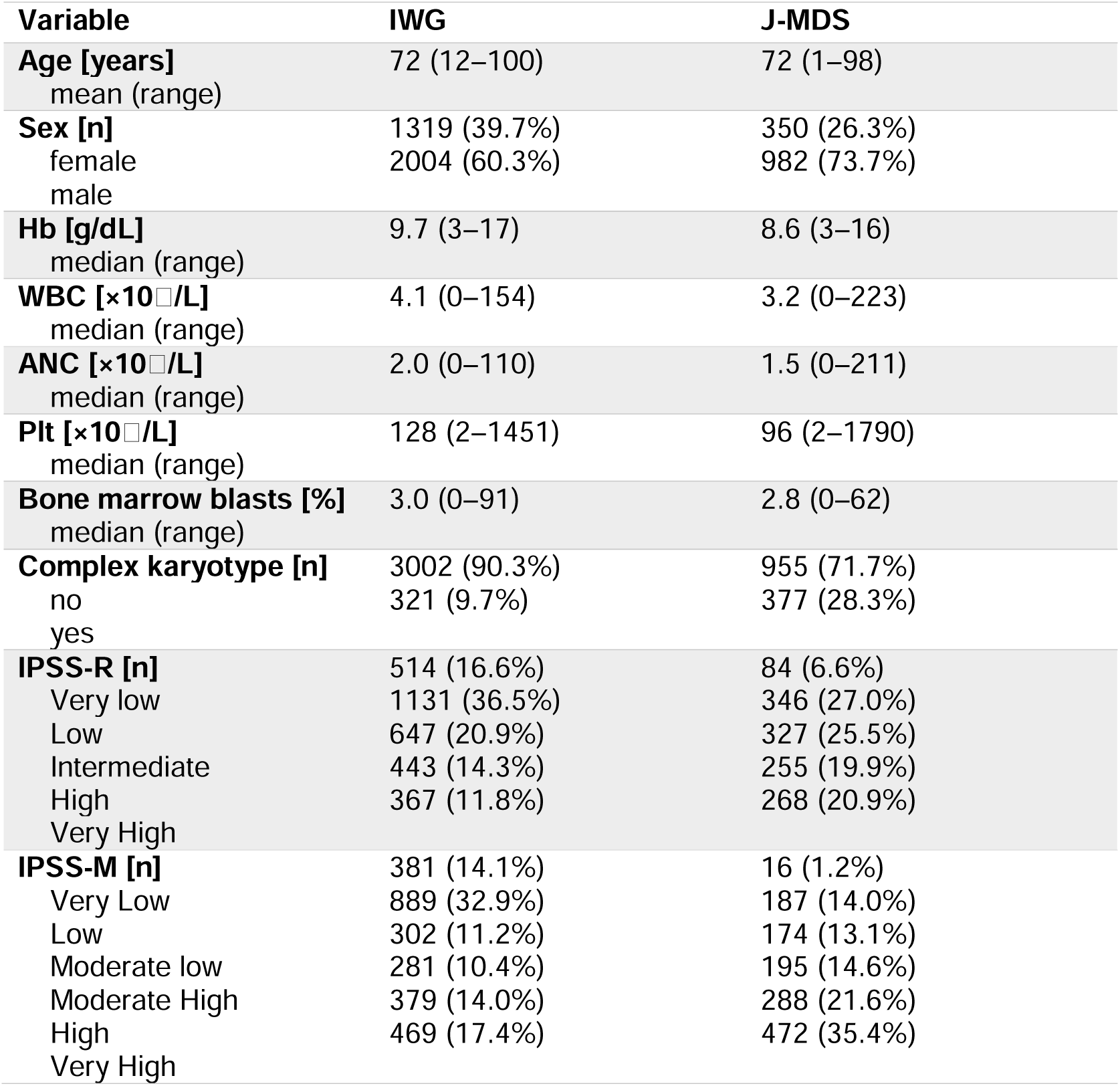
Patient characteristics between the cohorts. The Japanese cohort shows overall a higher risk composition as previously observed [2].

**Table S2.** Patient characteristics between PS-low and PS-high patients.

| **Variable** | **PS-low (<7)** | **PS-high (≥7)** |
| --- | --- | --- |
| **Age [years]** |  |  |
| median (range) | 74 (32–94) | 72 (21–92) |
| **Sex [n]** |  |  |
| female | 53 (51.5%) | 35 (33.0%) |
| male | 50 (48.5%) | 71 (67.0%) |
| **Hb [g/dL]** |  |  |
| median (range) | 9.2 (4–15) | 9.3 (4–15) |
| **WBC [$\times 10^9$/L]** |  |  |
| median (range) | 3.7 (1–134) | 3.2 (1–17) |
| **ANC [$\times 10^9$/L]** |  |  |
| median (range) | 1.9 (0–20) | 1.7 (0–13) |
| **Plt [$\times 10^9$/L]** |  |  |
| median (range) | 133.0 (5–1451) | 92.5 (9–906) |
| **Bone marrow blasts [%]** |  |  |
| median (range) | 3.0 (0–73) | 4.2 (0–36) |
| **Complex karyotype [n]** |  |  |
| no | 74 (71.8%) | 71 (67.0%) |
| yes | 29 (28.2%) | 35 (33.0%) |
| **IPSS-R [n]** |  |  |
| Very low | 7 (7.1%) | 12 (12.1%) |
| Low | 30 (30.3%) | 24 (24.2%) |
| Intermediate | 24 (24.2%) | 21 (21.2%) |
| High | 24 (24.2%) | 20 (20.2%) |
| Very High | 14 (14.1%) | 22 (22.2%) |
| **IPSS-M [n]** |  |  |
| Very Low | 6 (6.7%) | 6 (6.4%) |
| Low | 23 (25.6%) | 19 (20.2%) |
| Moderate low | 9 (10.0%) | 15 (16.0%) |
| Moderate High | 16 (17.8%) | 10 (10.6%) |
| High | 22 (24.4%) | 27 (28.7%) |
| Very High | 14 (15.6%) | 17 (18.1%) |

**Supplementary Table 1.** List of all detected monoallelic *TP53* (*TP53^mono^*) variants.

List of all detected monoallelic *TP53* (*TP53<sup>mono</sup>*) variants
| Variant | Variant Classification | No. of cases |
| --- | --- | --- |
| p.R175H | Missense_Mutation | 12 (5.1%) |
| p.R273H | Missense_Mutation | 12 (5.1%) |
| p.R248Q | Missense_Mutation | 9 (3.8%) |
| p.R273C | Missense_Mutation | 7 (3%) |
| p.V272M | Missense_Mutation | 5 (2.1%) |
| p.V216M | Missense_Mutation | 4 (1.7%) |
| p.G245S | Missense_Mutation | 3 (1.3%) |
| p.H193R | Missense_Mutation | 3 (1.3%) |
| p.R248W | Missense_Mutation | 3 (1.3%) |
| p.V157F | Missense_Mutation | 3 (1.3%) |
| p.Y220C | Missense_Mutation | 3 (1.3%) |
| p.C141Y | Missense_Mutation | 2 (0.8%) |
| p.C176S | Missense_Mutation | 2 (0.8%) |
| p.C238Y | Missense_Mutation | 2 (0.8%) |
| p.C277Y | Missense_Mutation | 2 (0.8%) |
| p.G244D | Missense_Mutation | 2 (0.8%) |
| p.G244S | Missense_Mutation | 2 (0.8%) |
| p.G245D | Missense_Mutation | 2 (0.8%) |
| p.H179D | Missense_Mutation | 2 (0.8%) |
| p.H179Q | Missense_Mutation | 2 (0.8%) |
| p.H179R | Missense_Mutation | 2 (0.8%) |
| p.H193L | Missense_Mutation | 2 (0.8%) |
| p.H214R | Missense_Mutation | 2 (0.8%) |
| p.I195S | Missense_Mutation | 2 (0.8%) |
| p.N235S | Missense_Mutation | 2 (0.8%) |
| p.P222L | Missense_Mutation | 2 (0.8%) |
| p.P250L | Missense_Mutation | 2 (0.8%) |
| p.R267W | Missense_Mutation | 2 (0.8%) |
| p.R273G | Missense_Mutation | 2 (0.8%) |
| p.R283C | Missense_Mutation | 2 (0.8%) |
| p.R337C | Missense_Mutation | 2 (0.8%) |
| p.S215G | Missense_Mutation | 2 (0.8%) |
| p.V172F | Missense_Mutation | 2 (0.8%) |
| p.Y163C | Missense_Mutation | 2 (0.8%) |
| p.Y220H | Missense_Mutation | 2 (0.8%) |
| p.Y234H | Missense_Mutation | 2 (0.8%) |
| p.Y234N | Missense_Mutation | 2 (0.8%) |
| c.375+1G>A | Splice_Site | 2 (0.8%) |
| p.C135fs | Frame_Shift_Del | 1 (0.4%) |
| p.C182fs | Frame_Shift_Del | 1 (0.4%) |
| p.E171fs | Frame_Shift_Del | 1 (0.4%) |
| p.N30Tfs*14 | Frame_Shift_Del | 1 (0.4%) |
| **p.P390Lfs\*32** | Frame_Shift_Del | 1 (0.4%) |
| **p.Y327fs** | Frame_Shift_Del | 1 (0.4%) |
| **p.I162Hfs\*19** | Frame_Shift_Ins | 1 (0.4%) |
| **p.K291Efs\*18** | Frame_Shift_Ins | 1 (0.4%) |
| **p.V73fs** | Frame_Shift_Ins | 1 (0.4%) |
| **p.I255del** | In_Frame_Del | 1 (0.4%) |
| **p.P177_C182del** | In_Frame_Del | 1 (0.4%) |
| **p.A159P** | Missense_Mutation | 1 (0.4%) |
| **p.A159V** | Missense_Mutation | 1 (0.4%) |
| **p.A161T** | Missense_Mutation | 1 (0.4%) |
| **p.A79T** | Missense_Mutation | 1 (0.4%) |
| **p.C176F** | Missense_Mutation | 1 (0.4%) |
| **p.C238G** | Missense_Mutation | 1 (0.4%) |
| **p.C275F** | Missense_Mutation | 1 (0.4%) |
| **p.C275R** | Missense_Mutation | 1 (0.4%) |
| **p.C275W** | Missense_Mutation | 1 (0.4%) |
| **p.C275Y** | Missense_Mutation | 1 (0.4%) |
| **p.D281G** | Missense_Mutation | 1 (0.4%) |
| **p.D281V** | Missense_Mutation | 1 (0.4%) |
| **p.D49N** | Missense_Mutation | 1 (0.4%) |
| **p.E224D** | Missense_Mutation | 1 (0.4%) |
| **p.E286V** | Missense_Mutation | 1 (0.4%) |
| **p.E287K** | Missense_Mutation | 1 (0.4%) |
| **p.F113V** | Missense_Mutation | 1 (0.4%) |
| **p.G244R** | Missense_Mutation | 1 (0.4%) |
| **p.G245V** | Missense_Mutation | 1 (0.4%) |
| **p.G266V** | Missense_Mutation | 1 (0.4%) |
| **p.G360V** | Missense_Mutation | 1 (0.4%) |
| **p.H178D** | Missense_Mutation | 1 (0.4%) |
| **p.H179N** | Missense_Mutation | 1 (0.4%) |
| **p.H179Y** | Missense_Mutation | 1 (0.4%) |
| **p.H193D** | Missense_Mutation | 1 (0.4%) |
| **p.I162F** | Missense_Mutation | 1 (0.4%) |
| **p.I195F** | Missense_Mutation | 1 (0.4%) |
| **p.I232S** | Missense_Mutation | 1 (0.4%) |
| **p.I254S** | Missense_Mutation | 1 (0.4%) |
| **p.I255T** | Missense_Mutation | 1 (0.4%) |
| **p.K132Q** | Missense_Mutation | 1 (0.4%) |
| **p.K132R** | Missense_Mutation | 1 (0.4%) |
| **p.K132T** | Missense_Mutation | 1 (0.4%) |
| **p.L111R** | Missense_Mutation | 1 (0.4%) |
| **p.L130V** | Missense_Mutation | 1 (0.4%) |
| **p.L137Q** | Missense_Mutation | 1 (0.4%) |
| **p.L194P** | Missense_Mutation | 1 (0.4%) |
| **p.L252P** | Missense_Mutation | 1 (0.4%) |
| **p.L257P** | Missense_Mutation | 1 (0.4%) |
| **p.L257R** | Missense_Mutation | 1 (0.4%) |
| **p.L265P** | Missense_Mutation | 1 (0.4%) |
| **p.L265R** | Missense_Mutation | 1 (0.4%) |
| **p.M133T** | Missense_Mutation | 1 (0.4%) |
| **p.M237I** | Missense_Mutation | 1 (0.4%) |
| **p.M237V** | Missense_Mutation | 1 (0.4%) |
| **p.N235T** | Missense_Mutation | 1 (0.4%) |
| **p.P151R** | Missense_Mutation | 1 (0.4%) |
| **p.P151S** | Missense_Mutation | 1 (0.4%) |
| **p.P177R** | Missense_Mutation | 1 (0.4%) |
| **p.P278A** | Missense_Mutation | 1 (0.4%) |
| **p.P278R** | Missense_Mutation | 1 (0.4%) |
| **p.P278S** | Missense_Mutation | 1 (0.4%) |
| **p.P98S** | Missense_Mutation | 1 (0.4%) |
| **p.Q5R** | Missense_Mutation | 1 (0.4%) |
| **p.R110P** | Missense_Mutation | 1 (0.4%) |
| **p.R156P** | Missense_Mutation | 1 (0.4%) |
| **p.R174G** | Missense_Mutation | 1 (0.4%) |
| **p.R175L** | Missense_Mutation | 1 (0.4%) |
| **p.R181C** | Missense_Mutation | 1 (0.4%) |
| **p.R202C** | Missense_Mutation | 1 (0.4%) |
| **p.R213G** | Missense_Mutation | 1 (0.4%) |
| **p.R213P** | Missense_Mutation | 1 (0.4%) |
| **p.R248L** | Missense_Mutation | 1 (0.4%) |
| **p.R249S** | Missense_Mutation | 1 (0.4%) |
| **p.R249T** | Missense_Mutation | 1 (0.4%) |
| **p.R273P** | Missense_Mutation | 1 (0.4%) |
| **p.R280G** | Missense_Mutation | 1 (0.4%) |
| **p.R280S** | Missense_Mutation | 1 (0.4%) |
| **p.R280T** | Missense_Mutation | 1 (0.4%) |
| **p.R282W** | Missense_Mutation | 1 (0.4%) |
| **p.R335H** | Missense_Mutation | 1 (0.4%) |
| **p.R337H** | Missense_Mutation | 1 (0.4%) |
| **p.R337S** | Missense_Mutation | 1 (0.4%) |
| **p.S116C** | Missense_Mutation | 1 (0.4%) |
| **p.S127F** | Missense_Mutation | 1 (0.4%) |
| **p.S241C** | Missense_Mutation | 1 (0.4%) |
| **p.S241F** | Missense_Mutation | 1 (0.4%) |
| **p.S241Y** | Missense_Mutation | 1 (0.4%) |
| **p.T155N** | Missense_Mutation | 1 (0.4%) |
| **p.T170M** | Missense_Mutation | 1 (0.4%) |
| **p.T256A** | Missense_Mutation | 1 (0.4%) |
| **p.T312S** | Missense_Mutation | 1 (0.4%) |
| **p.V143M** | Missense_Mutation | 1 (0.4%) |
| **p.V173M** | Missense_Mutation | 1 (0.4%) |
| **p.V197M** | Missense_Mutation | 1 (0.4%) |
| **p.V203M** | Missense_Mutation | 1 (0.4%) |
| **p.V272E** | Missense_Mutation | 1 (0.4%) |
| **p.V274L** | Missense_Mutation | 1 (0.4%) |
| **p.Y234D** | Missense_Mutation | 1 (0.4%) |
| **p.C277\*** | Nonsense_Mutation | 1 (0.4%) |
| **p.E343\*** | Nonsense_Mutation | 1 (0.4%) |
| **p.E68\*** | Nonsense_Mutation | 1 (0.4%) |
| **p.Q104\*** | Nonsense_Mutation | 1 (0.4%) |
| **p.R213\*** | Nonsense_Mutation | 1 (0.4%) |
| **p.R306\*** | Nonsense_Mutation | 1 (0.4%) |
| **p.R342\*** | Nonsense_Mutation | 1 (0.4%) |
| **p.S166\*** | Nonsense_Mutation | 1 (0.4%) |
| **p.S94\*** | Nonsense_Mutation | 1 (0.4%) |
| **p.Y103\*** | Nonsense_Mutation | 1 (0.4%) |
| **p.Y107_G108delins\*** | Nonsense_Mutation | 1 (0.4%) |
| **c.376-1G>A** | Splice_Site | 1 (0.4%) |
| **c.376-1G>T** | Splice_Site | 1 (0.4%) |
| **c.559+2T>G** | Splice_Site | 1 (0.4%) |
| **c.560-1G>A** | Splice_Site | 1 (0.4%) |
| **c.672+1G>T** | Splice_Site | 1 (0.4%) |
| **c.783-1G>A** | Splice_Site | 1 (0.4%) |
| **c.919+1G>A** | Splice_Site | 1 (0.4%) |

**Supplementary Table 2.**
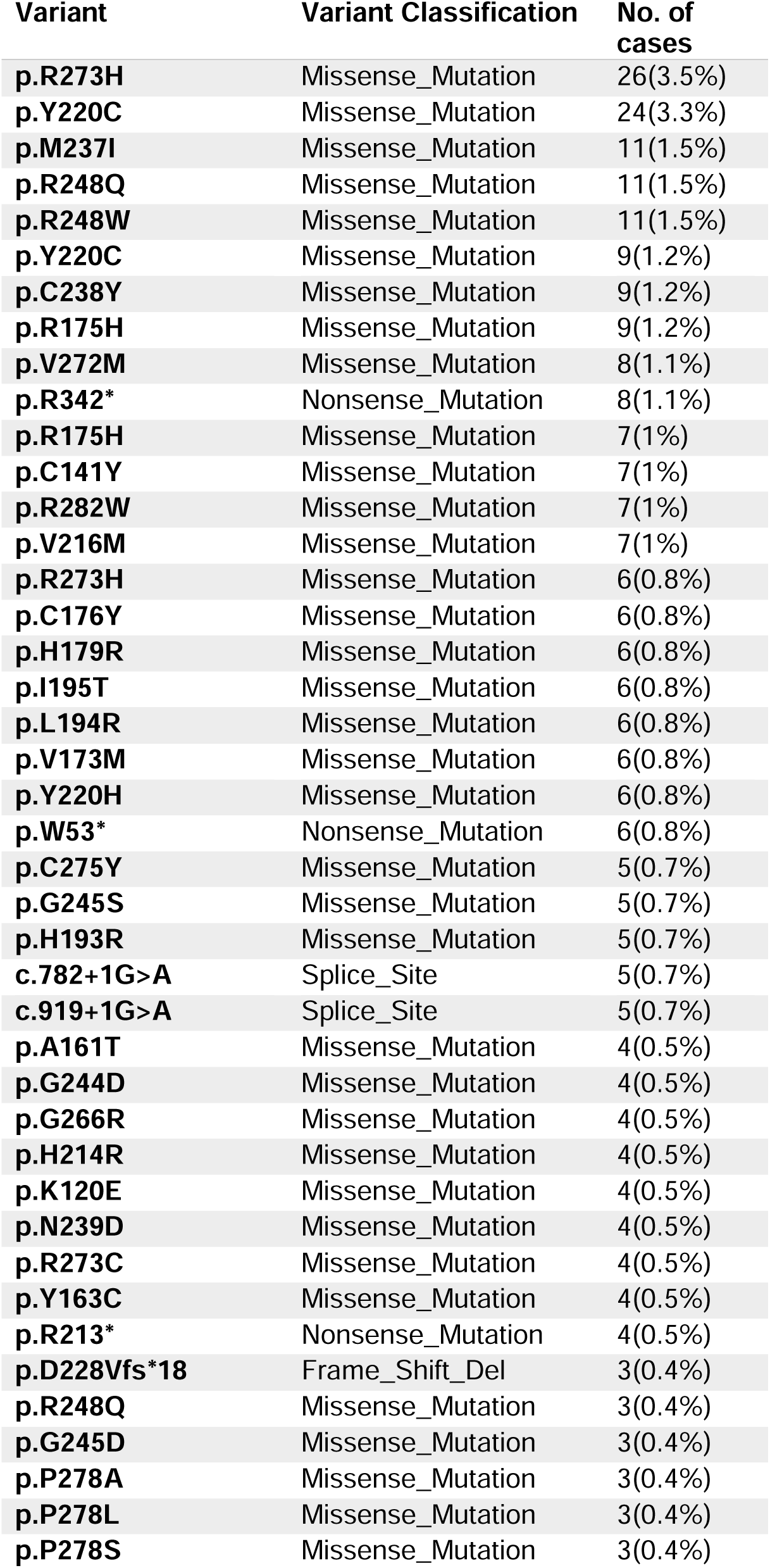
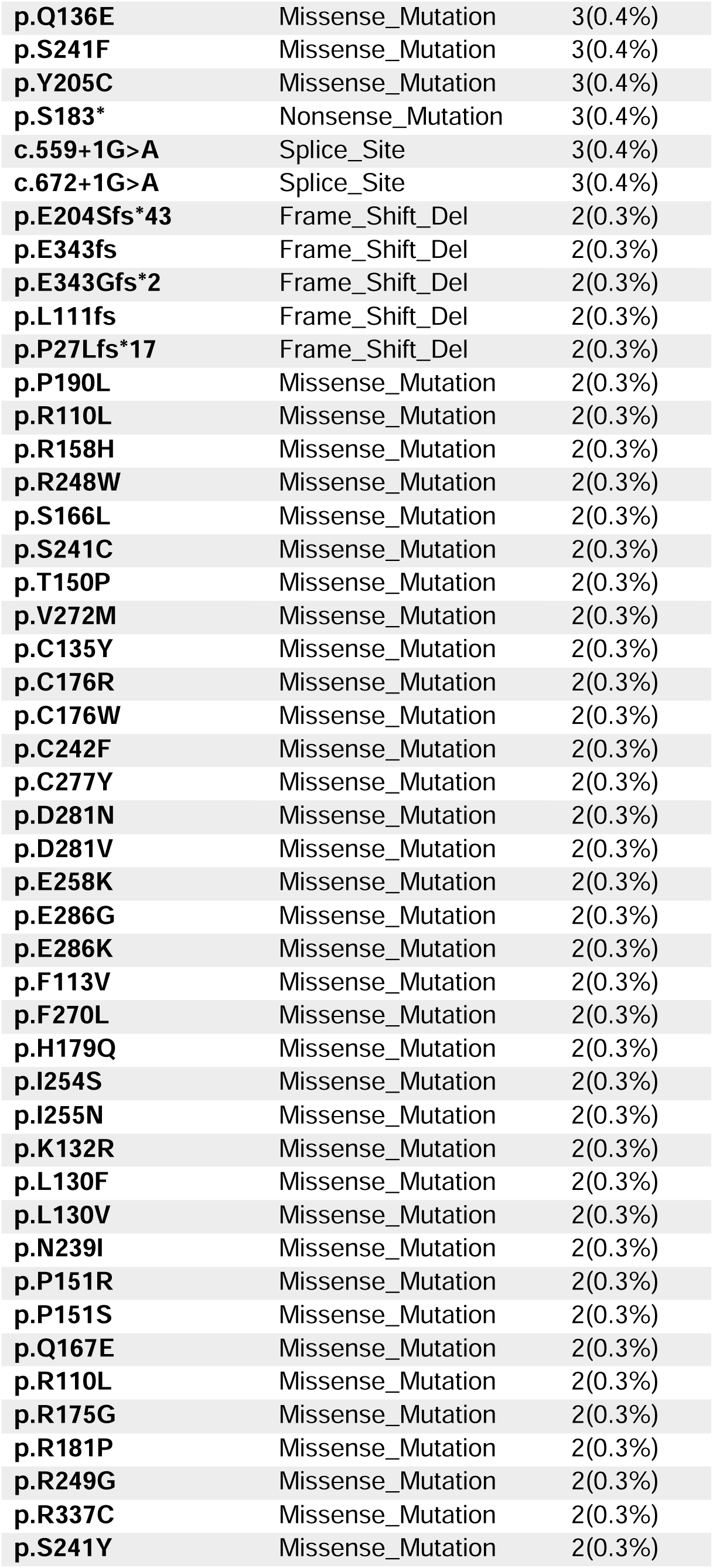
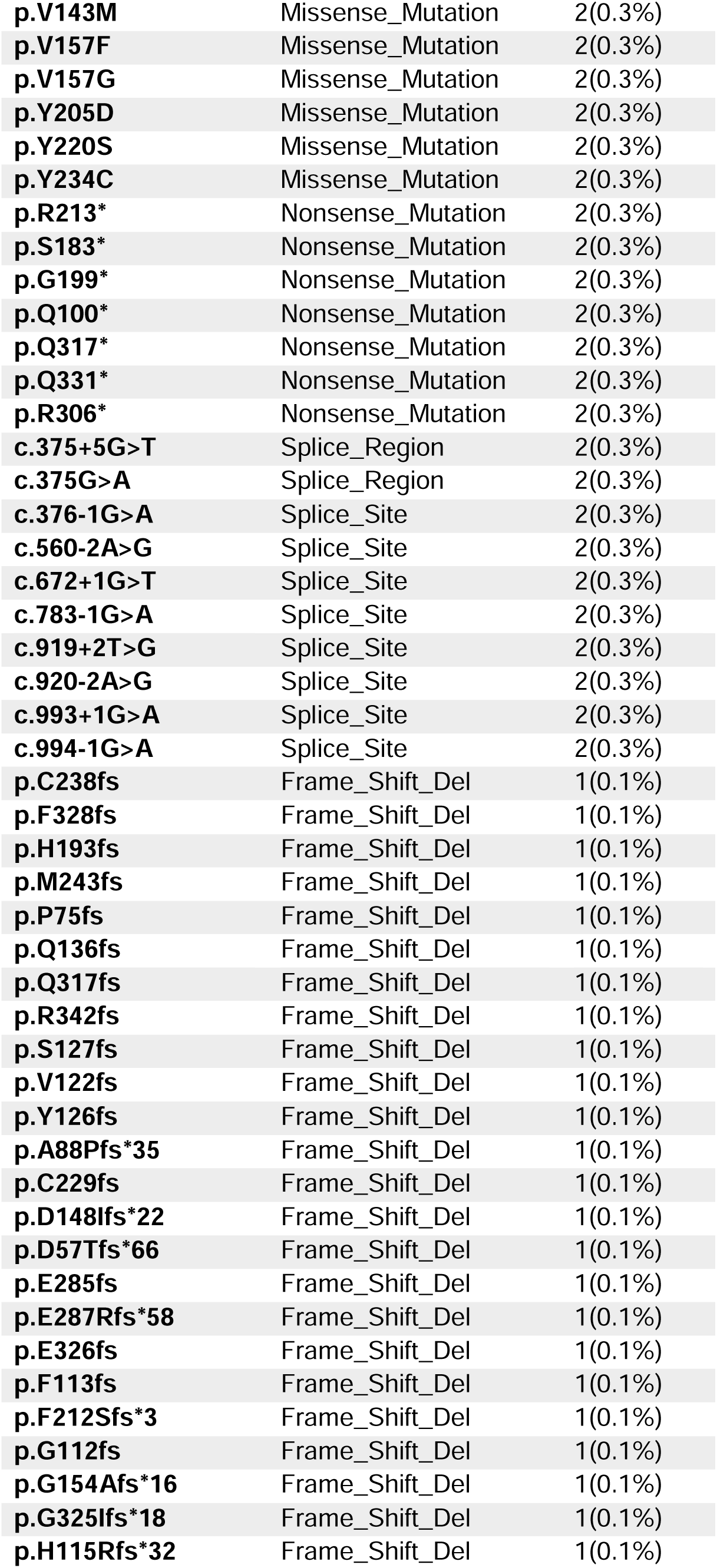
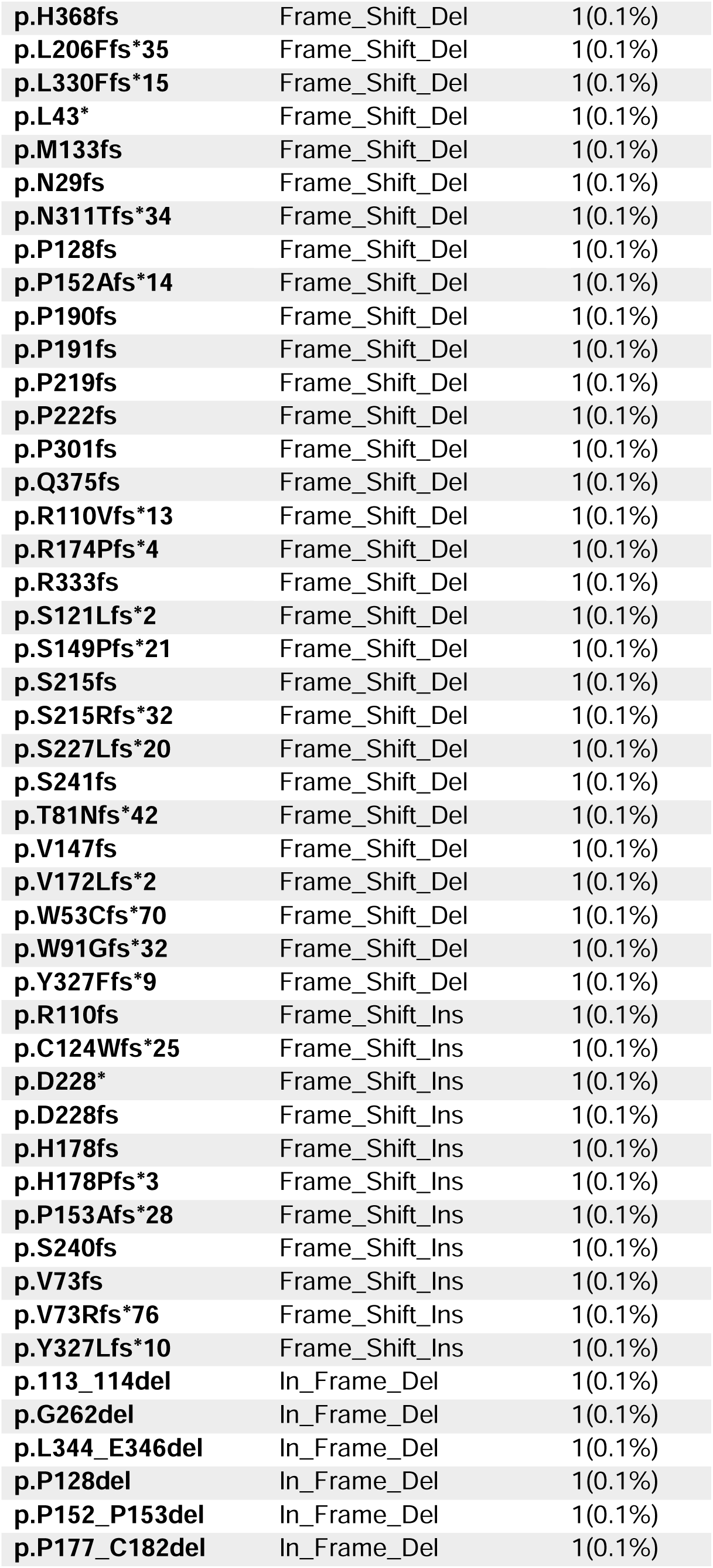
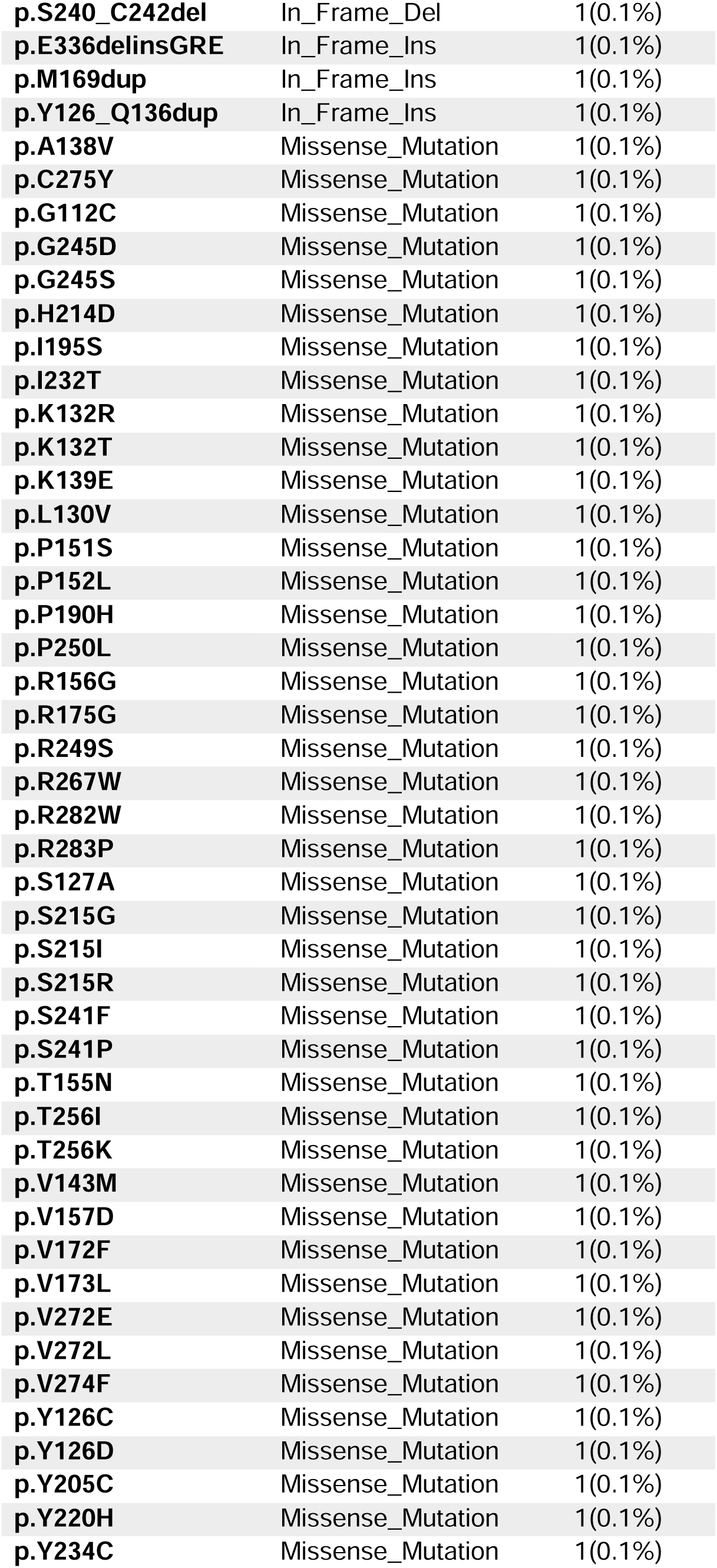
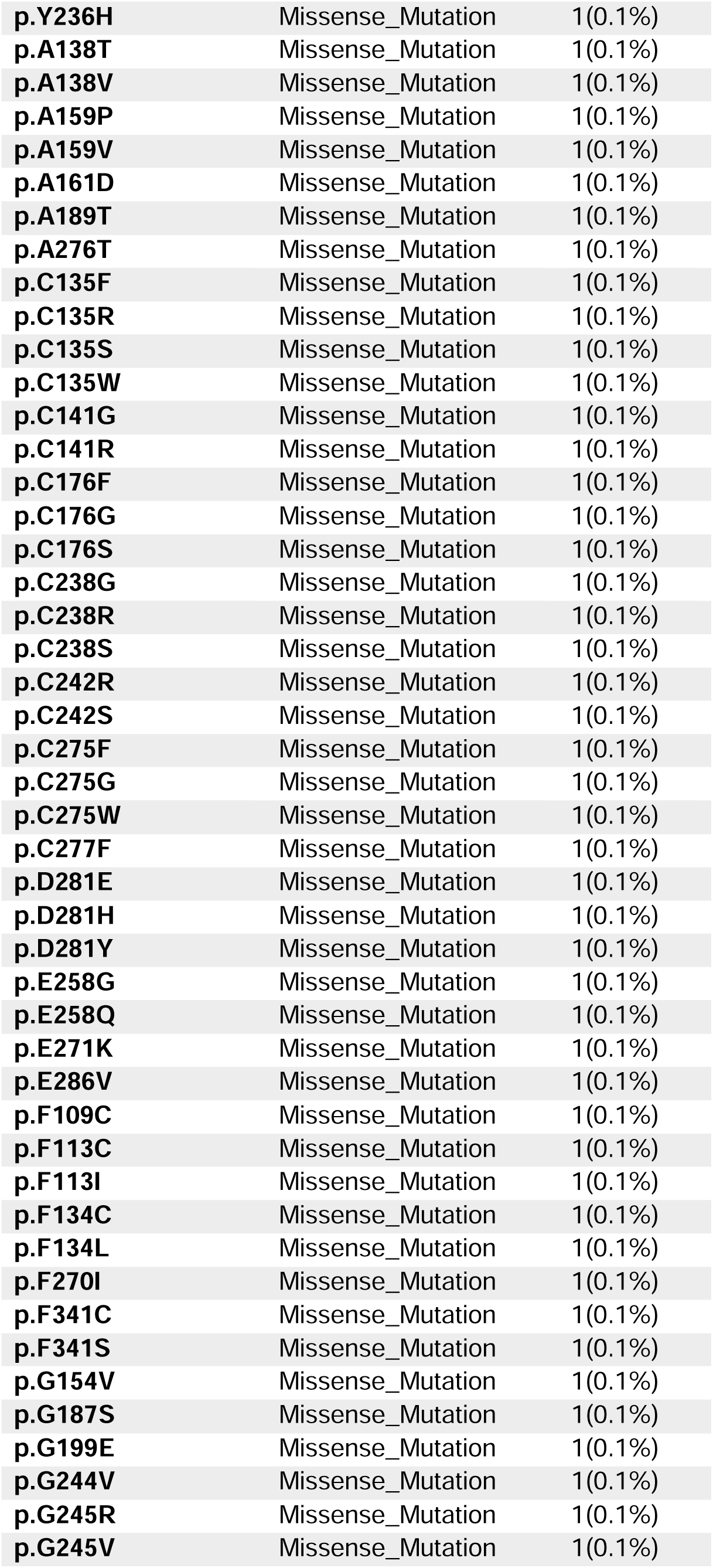
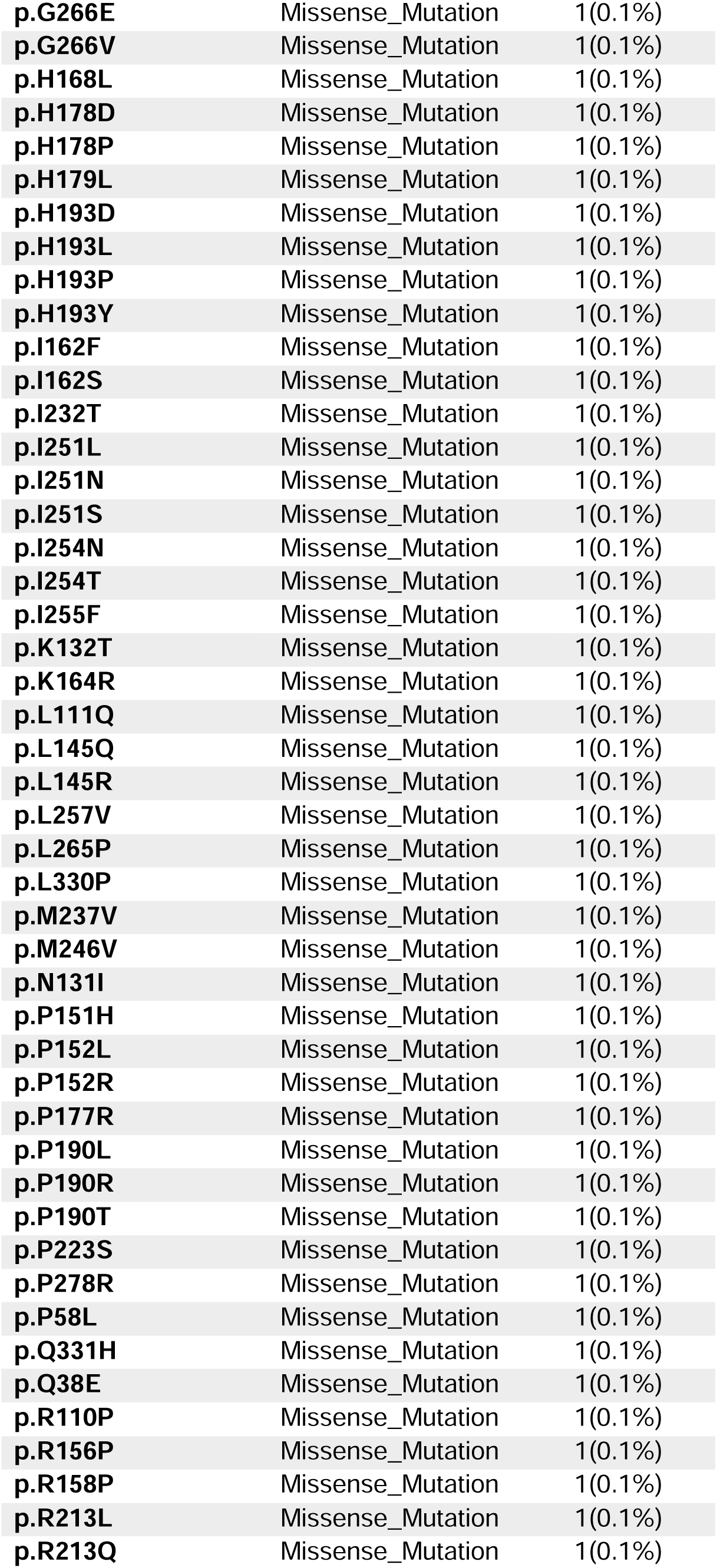
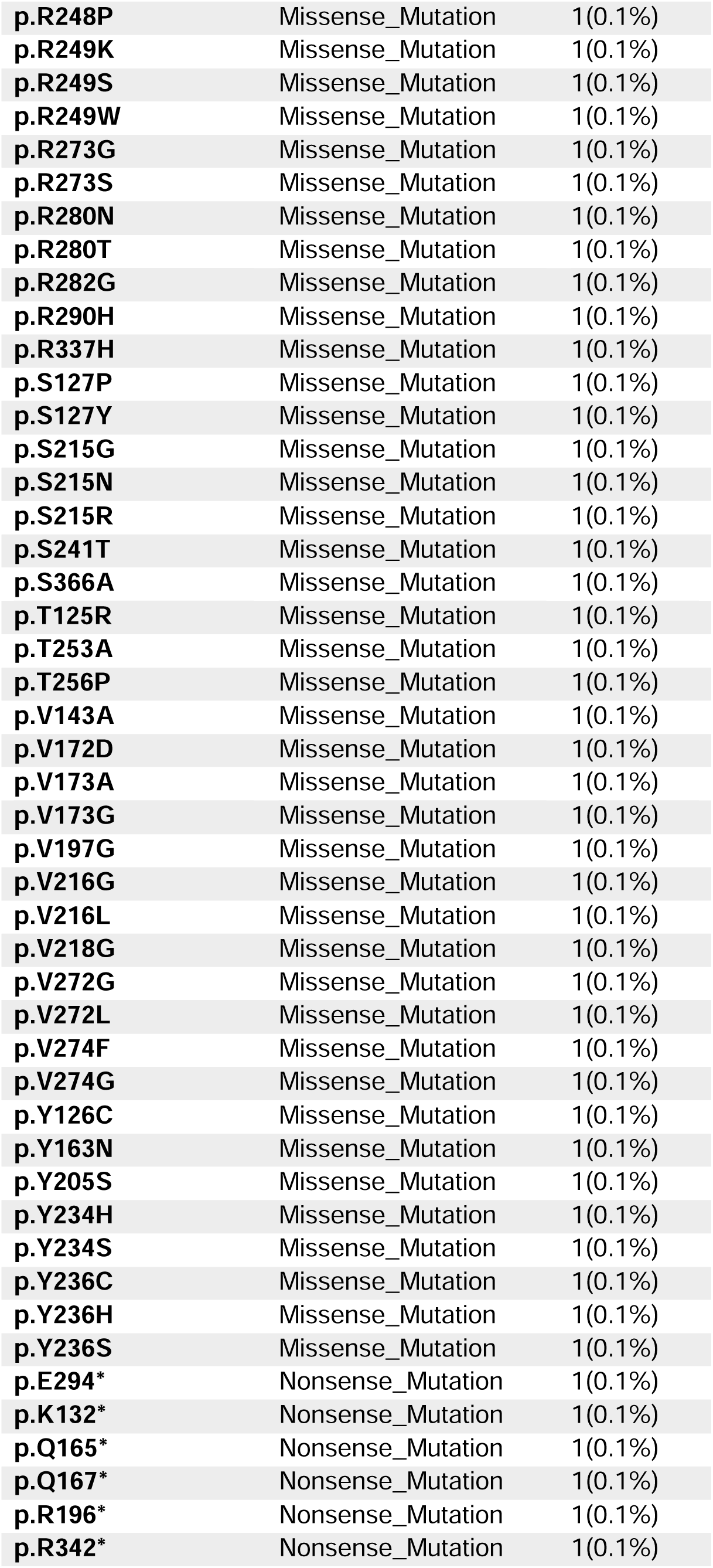
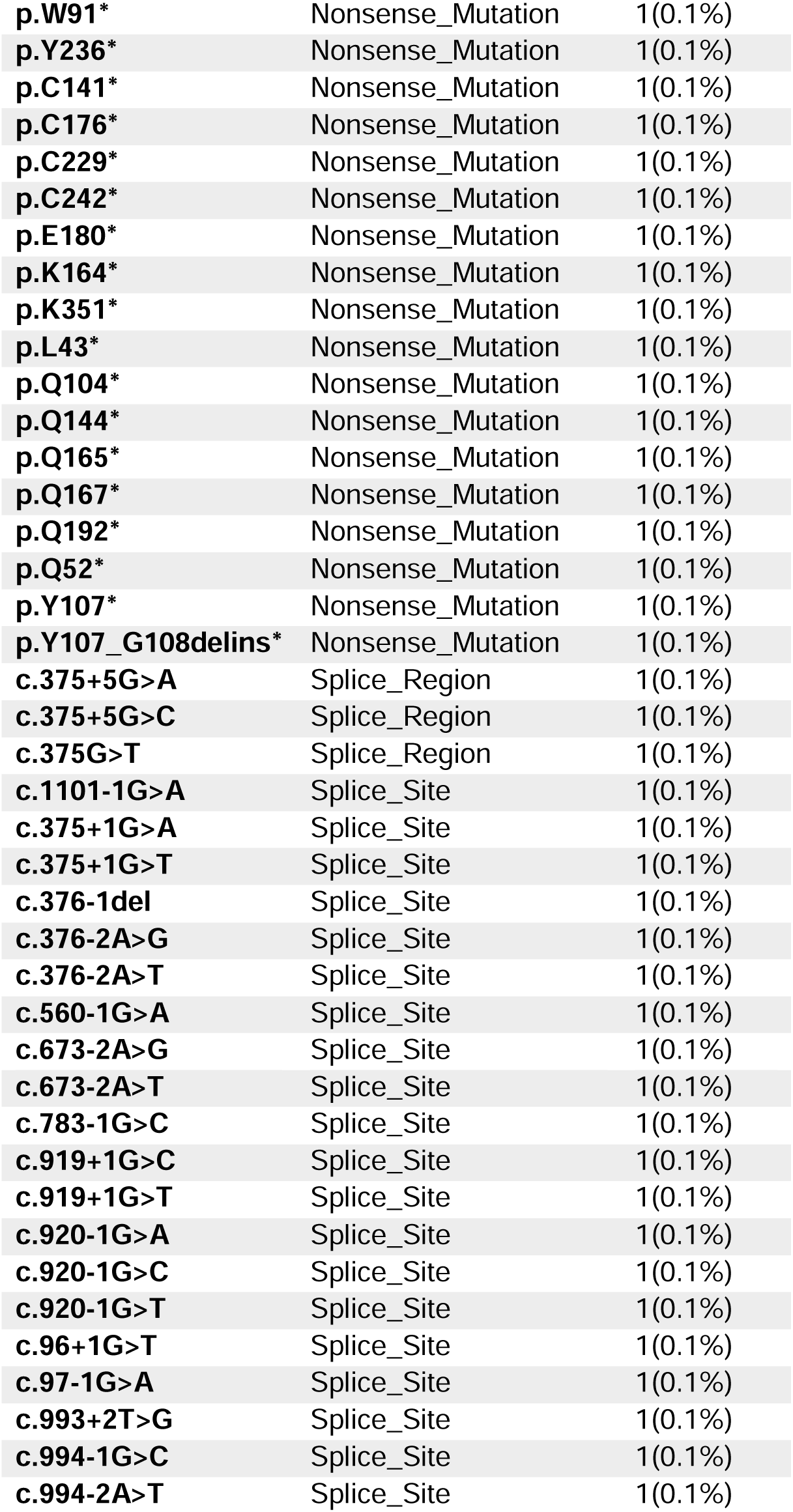
List of all detected multi-Hit *TP53* (*TP53^multiHit^*) variants.

## References

1. Khoury, J.D., et al., The 5th edition of the World Health Organization Classification of Haematolymphoid Tumours: Myeloid and Histiocytic/Dendritic Neoplasms. Leukemia, 2022. 36(7): p. 1703–1719.

2. Bernard, E., et al., Molecular International Prognostic Scoring System for Myelodysplastic Syndromes. NEJM Evidence, 2022. 1(7): p. EVIDoa2200008.

3. Boettcher, S., et al., A dominant-negative effect drives selection of TP53 missense mutations in myeloid malignancies. Science, 2019. 365(6453): p. 599–604.

4. Lindsley, R.C., et al., Prognostic Mutations in Myelodysplastic Syndrome after Stem-Cell Transplantation. N Engl J Med, 2017. 376(6): p. 536–547.

5. Bernard, E., et al., Implications of TP53 allelic state for genome stability, clinical presentation and outcomes in myelodysplastic syndromes. Nature Medicine, 2020. 26(10): p. 1549–1556.

6. Haase, D., et al., TP53 mutation status divides myelodysplastic syndromes with complex karyotypes into distinct prognostic subgroups. Leukemia, 2019. 33(7): p. 1747–1758.

7. Montoro, M.J., et al., Influence of TP53 gene mutations and their allelic status in myelodysplastic syndromes with isolated 5q deletion. Blood, 2024. 144(16): p. 1722–1731.

8. Sallman, D.A., et al., Impact of TP53 mutation variant allele frequency on phenotype and outcomes in myelodysplastic syndromes. Leukemia, 2016. 30(3): p. 666–673.

9. Wang, W., et al., Characterization of TP53 mutations in low-grade myelodysplastic syndromes and myelodysplastic syndromes with a non-complex karyotype. Eur J Haematol, 2017. 99(6): p. 536–543.

10. Weinberg, O.K., et al., TP53 mutation defines a unique subgroup within complex karyotype de novo and therapy-related MDS/AML. Blood Adv, 2022. 6(9): p. 2847–2853.

11. Vogelstein, B., D. Lane, and A.J. Levine, Surfing the p53 network. Nature, 2000. 408(6810): p. 307–310.

12. Milner, J. and E.A. Medcalf, Cotranslation of activated mutant p53 with wild type drives the wild-type p53 protein into the mutant conformation. Cell, 1991. 65(5): p. 765–74.

13. Vousden, K.H. and D.P. Lane, p53 in health and disease. Nat Rev Mol Cell Biol, 2007. 8(4): p. 275–83.

14. Aubrey, B.J., A. Strasser, and G.L. Kelly, Tumor-Suppressor Functions of the TP53 Pathway. Cold Spring Harb Perspect Med, 2016. 6(5).

15. Wang, H., et al., Targeting p53 pathways: mechanisms, structures, and advances in therapy. Signal Transduction and Targeted Therapy, 2023. 8(1): p. 92.

16. Ayaz, G., et al., An Updated View of the Roles of p53 in Embryonic Stem Cells. Stem Cells, 2022. 40(10): p. 883–891.

17. Jambhekar, A., et al., Comparison of TP53 mutations in myelodysplasia and acute leukemia suggests divergent roles in initiation and progression. Blood Neoplasia, 2024. 1(1).

18. Vaddavalli, P.L. and B. Schumacher, The p53 network: cellular and systemic DNA damage responses in cancer and aging. Trends in Genetics, 2022. 38(6): p. 598–612.

19. Trbusek, M., et al., Missense Mutations Located in Structural p53 DNA-Binding Motifs Are Associated With Extremely Poor Survival in Chronic Lymphocytic Leukemia. Journal of Clinical Oncology, 2011. 29(19): p. 2703–2708.

20. Román-Rosales, A.A., et al., Mutant p53 gain of function induces HER2 over-expression in cancer cells. BMC Cancer, 2018. 18(1): p. 709.

21. Stein, Y., V. Rotter, and R. Aloni-Grinstein, Gain-of-Function Mutant p53: All the Roads Lead to Tumorigenesis. Int J Mol Sci, 2019. 20(24).

22. Giacomelli, A.O., et al., Mutational processes shape the landscape of TP53 mutations in human cancer. Nature Genetics, 2018. 50(10): p. 1381–1387.

23. Kataoka, K., et al., Integrated molecular analysis of adult T cell leukemia/lymphoma. Nat Genet, 2015. 47(11): p. 1304–15.

24. Yoshizato, T., et al., Genetic abnormalities in myelodysplasia and secondary acute myeloid leukemia: impact on outcome of stem cell transplantation. Blood, 2017. 129(17): p. 2347–2358.

25. Santini, V., M. Stahl, and D.A. Sallman, TP53 Mutations in Acute Leukemias and Myelodysplastic Syndromes: Insights and Treatment Updates. American Society of Clinical Oncology Educational Book, 2024. 44(3): p. e432650.

26. Shin, D.Y., TP53 Mutation in Acute Myeloid Leukemia: An Old Foe Revisited. Cancers (Basel), 2023. 15(19).

27. George, B., et al., TP53 in Acute Myeloid Leukemia: Molecular Aspects and Patterns of Mutation. International Journal of Molecular Sciences, 2021. 22(19): p. 10782.

28. Arber, D.A., et al., International Consensus Classification of Myeloid Neoplasms and Acute Leukemias: integrating morphologic, clinical, and genomic data. Blood, 2022. 140(11): p. 1200–1228.

29. Lausen, B. and M. Schumacher, Maximally Selected Rank Statistics. Biometrics, 1992. 48(1): p. 73–85.

